# A Transformer-Based 2.5D Deep Learning Model for Preoperative Prediction of Lymph Node Metastasis in Papillary Thyroid Carcinoma

**DOI:** 10.64898/2026.04.01.26349933

**Authors:** Shaojie Xu, Xingmin Yan, Yuhang Su, Jia Qi, Xingyu Chen, Yulin Li, Hao Xiong, Jiamei Jiang, Zhihao Wei, Ziying Chen, Yisikandaer Yalikun, Hanning Li, Xingyin Li, Yiqing Xi, Wei Li, Xingrui Li, Yaying Du

**Affiliations:** Department of Thyroid and Breast Surgery, Tongji Hospital, Tongji Medical College, Huazhong University of Science and Technology, Wuhan, Hubei, China; Division of Hepato-Pancreato-Biliary Surgery, Tongji Hospital, Tongji Medical College, Huazhong University of Science and Technology, Wuhan, Hubei, China; Department of Thyroid and Breast Surgery, The Third Affiliated Hospital of Sun Yat-Sen University, Guangzhou, China; Tongji Medical College, Huazhong University of Science and Technology, Wuhan, Hubei, China; Radiology Department, Nanchuan Hospital of Chongqing Medical University, Chongqing, China; Radiology Department, Red Cross Hospital of Yulin City, YuLin, Guangxi Zhuang Autonomous Region, China; Department of Physician Services, Xinjiang International Medical Center (Xinjiang Medical Hospital), Urumqi, China; Department of Head and Neck Surgery, Hubei Cancer Hospital, Tongji Medical College, Huazhong University of Science and Technology, Wuhan, Hubei, China; Department of Nuclear Medicine, Tianjin Medical University General Hospital, Tianjin, China

**Keywords:** Transformer-based model, Lymph node metastasis, Papillary thyroid carcinoma, Deep learning in medical imaging, Preoperative prediction. These findings suggest that ThyLNT provides robust predictive performance while capturing biologically relevant features of metastatic progression

## Abstract

**Background:** Accurate preoperative prediction of lymph node metastasis (LNM) in papillary thyroid carcinoma (PTC) remains challenging, particularly in clinically node-negative (cN0) patients, leading to potential overtreatment. We aimed to develop and validate a Transformer-based 2.5D deep learning model (ThyLNT) using preoperative computed tomography (CT) images for robust prediction of LNM and to explore its underlying biological basis through multi-omics analyses.

**Methods:** A total of 1,560 PTC patients from six hospitals were retrospectively included. The Tongji Hospital cohort (n=1,010) was divided into training (70%) and internal validation (30%) sets, while five independent institutions served as external test cohorts. For each lesion, seven 2.5D slices were extracted and modeled using a DenseNet201 backbone. Slice-level features were integrated using a Transformer-based feature-level fusion strategy and compared with ensemble learning, multi-instance learning (MIL), and traditional radiomics approaches. Model performance was assessed using area under the receiver operating characteristic curve (AUC), calibration analysis, decision curve analysis (DCA), and precision-recall curves. Multi-omics analyses, including bulk RNA-seq, single-cell RNA-seq, spatial transcriptomics, and spatial metabolomics, were performed to investigate biological correlates.

**Results:** The Transformer-based model consistently outperformed comparator models across cohorts. In the training and validation cohorts, ThyLNT achieved AUCs of 0.882 and 0.787, respectively, with external AUCs ranging from 0.772 to 0.827. Compared with ultrasound (US) and CT, ThyLNT showed superior predictive performance (all P < 0.001 in the validation cohort). Simulation analysis in cN0 patients suggested that ThyLNT could reduce unnecessary lymph node dissection (LND) from 52.16% to 4.88%. Transcriptomic analysis combined with WGCNA and correlation analysis identified VEGFA as the gene most strongly associated with ThyLNT prediction scores. Single-cell and spatial transcriptomic analyses suggested metastasis-related tumor microenvironment remodeling, while enrichment analysis of genes affected by virtual knockout of VEGFA indicated involvement of angiogenesis- and epithelial–mesenchymal transition (EMT)-related pathways. Spatial metabolomics further revealed coordinated lipid metabolic reprogramming in metastatic tissues. These findings suggest that ThyLNT provides robust predictive performance while capturing biologically relevant features associated with metastatic progression.

## Introduction

Papillary thyroid carcinoma (PTC) is the most prevalent thyroid malignancy, with a rapidly increasing incidence^1^. Approximately 30% 80% of patients with PTC develop cervical lymph node metastasis (LNM) at an early stage^2^, including central lymph node metastasis (CLNM) and lateral lymph node metastasis (LLNM). Some patients even present initially with painless cervical lymphadenopathy as their first clinical manifestation^3^. Inadequate evaluation of lateral lymph nodes may result in early recurrence and reduced overall survival^4^. Improving the diagnostic accuracy of cervical LNM not only helps optimize risk stratification and surgical decision-making but may also reduce unnecessary lymph node dissection (LND), thereby decreasing surgical complications and healthcare costs.

Cervical US is the first-line preoperative assessment for PTC. Neck CT is commonly added to improve evaluation of central/lateral nodal compartments and surrounding anatomy, particularly in cases with limited sonographic windows or suspected extensive disease^5^. However, the assessment of cervical LNM in PTC using conventional imaging largely depends on physician experience, leading to substantial interobserver variability. Factors such as micrometastases, atypical morphology, and anatomical obstruction make certain metastatic lesions difficult to detect by preoperative US or CT^6^, resulting in a relatively high rate of missed diagnoses.

Artificial intelligence based on CT imaging has demonstrated diagnostic performance comparable to that of radiologists in detecting cervical LNM in patients with PTC^7^. Most existing deep learning approaches rely on 2D slice-based modeling, for example selecting the largest tumor slice as the representative image, thereby failing to capture spatial continuity across adjacent CT slices. Although fully 3D models can exploit volumetric information, they demand high computational resources and are prone to overfitting with limited medical data^8^. The 2.5D paradigm, which incorporates adjacent cross-sections to approximate volumetric context while maintaining manageable complexity, has emerged as a practical compromise. However, 2.5D modeling introduces a critical challenge: how to aggregate slice-level representations into a coherent prediction. Conventional fusion strategies, such as probability averaging or multiple instance learning (MIL), implicitly assume partial interchangeability among slices and do not explicitly model inter-slice dependency^9^. Given that metastatic propensity may manifest as a distributed imaging phenotype across multiple planes rather than being confined to a single dominant slice, effective cross-slice modeling becomes essential. Transformer architectures, through self-attention mechanisms that capture long-range relationships, provide a principled framework for feature-level integration across slices^10^.

Based on this rationale, the present study selected specific axial slices from the tumor regions of interest (ROI) on CT images and extracted deep features. A convolutional neural network (CNN) was first employed to obtain high-dimensional representation features from the penultimate layer, followed by feature-level fusion using a Transformer to integrate multi-slice information and generate patient-level predictions of metastatic risk.We further conducted systematic comparisons with fusion strategies such as ensemble learning and MIL to validate the advantages of Transformer-based fusion for this task. To elucidate the potential biological basis underlying the “black-box” model predictions, this study incorporated multi-omics analyses to explore molecular pathways and microenvironmental characteristics associated with predicted risk at the transcriptomic, single-cell, and spatial levels, thereby providing mechanistic support for the imaging-based predictions. The overall study workflow is illustrated in Figure 1.

**Figure 1.**
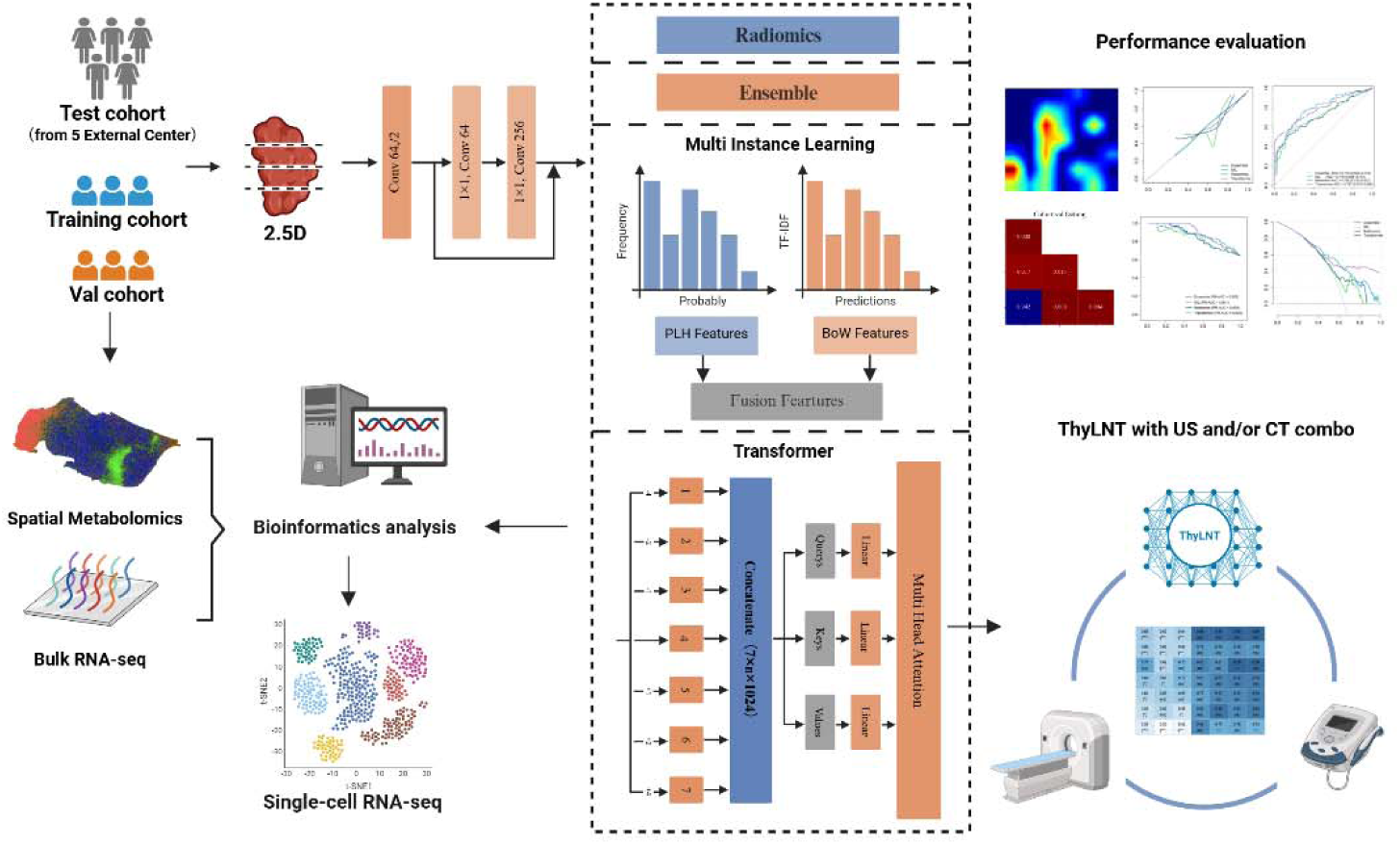
Schematic representation of the study design. Created in https://BioRender.com

## Methods

### Data Sets

Data for the development and validation of ThyLNT were retrospectively derived from six hospitals (Tongji Medcial college of Huazhong University of Science and Technology (TJH), Tianjin Medical University General Hospital (TMUGH), Xinjiang International Hospital (XIMC), Hubei Cancer Hospital (HBCH), the Third Affiliated Hospital of Sun Yat-sen University (SYSUTH) and Nanchuan hospital of Chongqing Medicial University (NCMU)) from January 2015 to December 2025. The inclusion criteria were as follows: (1) age ≥18 years; (2) pathologically confirmed PTC following thyroidectomy; (3) availability of preoperative thyroid ultrasonography and CT. The exclusion criteria were as follows: (1) presence of disease recurrence at admission; (2) evidence of distant metastasis; (3) incomplete preoperative thyroid ultrasonography or CT data; and (4) poor-quality thyroid CT images (e.g., severe artifacts or inadequate image clarity). The TJH dataset was randomly divided into two subsets: a training cohort comprising 70% of the cases and an internal validation cohort consisting of the remaining 30%. To evaluate the generalizability of the model, data from the other five institutions were used as independent external test cohorts. All personal identifiers were removed, and samples/datasets assigned unique anonymous codes.

### Image Segmentation and Preprocessing

The ROI were delineated using ITK-SNAP by two experienced radiologists independently. In cases of disagreement, a third radiologist with over 20 years of expertise resolved the discrepancies, ensuring the accuracy and reliability of the ROI annotations. Voxel intensities were standardized, with the window width and level adjusted to 50 and 350, respectively. CT images were resampled to a uniform voxel spacing of 1X 1X 1mmusing bilinear interpolation for the images and nearest-neighbor interpolation for the labels, ensuring consistency in resolution across the dataset.

### Data Preparation

For each lesion, 2.5D data cropping was performed to enhance the representation of the ROI. The process began by identifying the largest cross-sectional area within the 3D image data, followed by extracting adjacent slices both above and below the primary slice. Specifically, slices were sampled from the superior-inferior plane at intervals of ±1, ±2, and ±4 slices relative to the central slice. This resulted in a set of seven 2D slices per patient, capturing both the primary cross-sectional region and surrounding contextual information. All slices shared the same label as the patient, forming the basis for subsequent model training.

### Model Training

Each slice was treated as an independent sample for training, with labels shared across slices to ensure consistency. A CNN was employed to model these 2.5D slices, utilizing transfer learning techniques with architectures pre-trained on the ImageNet dataset, including DenseNet121, ResNet101, ResNet50, DenseNet201, and ResNet18. Model details are provided in Supplementary Material 1A.

### Fusion Model Development

To enhance predictive performance, we developed a Transformer-based model specifically designed for feature-level integration, enabling the aggregation of deep features extracted from all seven slices of each lesion. The detailed architecture of the Transformer model is presented in Supplementary Figure 1, with additional technical descriptions provided in Supplementary Material 2A. In parallel, alternative fusion strategies were also evaluated, including MIL (Supplementary Material 2B) and an ensemble approach (Supplementary Material 2C). In addition, a traditional radiomics methodology was implemented as a reference model (Supplementary Material 3A).

These approaches were comprehensively assessed to compare their effectiveness in aggregating multi-slice information and improving predictive performance. By systematically evaluating the strengths and limitations of each method, we sought to establish a robust and comprehensive framework for 2.5D data analysis.

The clinical assessment of LNM was extracted from the official radiology reports in the medical records. LNM was considered positive based on US or CT assessment if lymph nodes were described as “malignant,” “metastatic,” “abnormal,” or “suspicious” in the report. We calculated the diagnostic performance of US alone, CT alone, and their combination (US+CT) in the overall cohort. For the combined imaging assessment (US+CT), the result was defined as negative (cN0) only when both US and CT showed no evidence of LNM. If either modality was reported as positive for LNM, the combined imaging result was considered clinically positive for LNM (cN1). The performance of ThyLNT in combination with conventional preoperative imaging was further evaluated using a logistic regression model. Furthermore, we assessed whether ThyLNT could reduce the rate of unnecessary LND in cN0 patients, in whom prophylactic central LND is routinely performed in China.

### Metrics

For slice-level models, evaluation metrics were calculated for individual slices, and overall performance was primarily assessed using the area under the receiver operating characteristic curve (AUC). To avoid information leakage, comparative evaluations were performed exclusively on the validation set.

For fused feature analysis, the task shifted to patient-level prediction of LNM status, diagnostic performance was assessed by constructing receiver operating characteristic (ROC) curves. Calibration curves were generated to evaluate the agreement between predicted probabilities and observed outcomes, and the Hosmer-Lemeshow test was performed to assess calibration quality. Decision curve analysis (DCA) was conducted to quantify the clinical utility and net benefit of the predictive models. Precision-recall (PR) curves were additionally plotted to further assess model performance, particularly under class imbalance conditions. The area under the PR curve (average precision, AP) was calculated to provide complementary information to the ROC-based AUC.

### Multi-omics Bioinformatics Analysis

Thirty-five patients with PTC with available bulk RNA-seq data were included in this study. After standard normalization and preprocessing of TPM expression data, differential expression analyses were performed based on both model-predicted groups and true clinical groups (P < 0.05), followed by Gene Ontology (GO) and Kyoto Encyclopedia of Genes and Genomes (KEGG) enrichment analyses to validate the biological consistency of the model predictions at the molecular level. To investigate coordinated gene expression patterns, weighted gene co-expression network analysis (WGCNA) was conducted. A signed co-expression network was constructed using an appropriate soft-thresholding power, and genes were clustered into modules by hierarchical clustering. Module eigengenes were calculated and correlated with model prediction scores and clinicopathological traits to identify relevant modules and candidate hub genes.

Single-cell RNA sequencing data were obtained from the publicly available dataset GSE184362. Data preprocessing, including quality control, normalization, dimensionality reduction, and clustering, was performed using Seurat. Cell types were annotated based on canonical marker genes^11^. Pseudotime trajectory analysis was conducted using Monocle to infer cellular state transitions and evaluate dynamic gene expression changes. Cell-cell communication analysis was performed using CellChat to infer ligand-receptor interactions and to characterize signaling roles, including sender, receiver, and mediator cells within specific pathways. In addition, in silico gene knockout analysis was performed using scTenifoldKnk to evaluate the regulatory impact of VEGFA on gene networks derived from single-cell RNA-seq data.

Spatial transcriptomic data were obtained from the public dataset GSE250521. Data preprocessing, normalization, dimensionality reduction, and spatial visualization were performed using standard analytical workflows to assess spatial gene expression patterns and cell type distribution^12^.

Spatial metabolomics analysis was performed on three LNM cases and three non-metastatic cases that were correctly classified by the model. After peak detection, alignment, and normalization, orthogonal partial least squares discriminant analysis (OPLS-DA) was applied to identify metabolites with high variable importance in projection (VIP) scores. Differential metabolites were determined by statistical testing, followed by pathway enrichment and correlation network analyses.

### Statistical Analysis

The Shapiro-Wilk test was used to assess the normality of continuous variables. Normally distributed variables were compared using the independent-samples t test, whereas non-normally distributed variables were analyzed using the Mann-Whitney U test. Categorical variables were compared using the chi-square (χ²) test. Continuous variables are presented as mean ± standard deviation (SD), and categorical variables as counts (percentages).

For each diagnostic strategy, 2 × 2 contingency tables were constructed to calculate sensitivity, specificity, positive predictive value (PPV), and negative predictive value (NPV). The 95% confidence intervals (CIs) were calculated using the exact Clopper-Pearson method. Differences between AUCs were compared using the two-sided DeLong test.

All statistical analyses were performed using SAS software (version 9.4; SAS Institute, Cary, NC, USA), and a two-sided P value < 0.05 was considered statistically significant. Machine learning model development and training were conducted using Python (version 3.7.12) with scikit-learn (version 1.0.2) and Onekey (version 3.3.5). Deep learning experiments were implemented using MONAI (version 0.8.1) and PyTorch (version 1.8.1) on an NVIDIA RTX 4090 GPU.

## Results

After patient selection (Supplementary Figure 2), a total of 1,010 PTC patients from TJH were included. These patients were randomly divided into a training cohort (n = 706) and an independent internal validation cohort (n = 304). The external validation cohorts consisted of 179 PTC patients from TMUGH, 148 from XIMC, 77 from HBCH, 77 from NCMU, and 69 from SYSUTH. Baseline patient characteristics are summarized in Table 1.

**Table 1.**
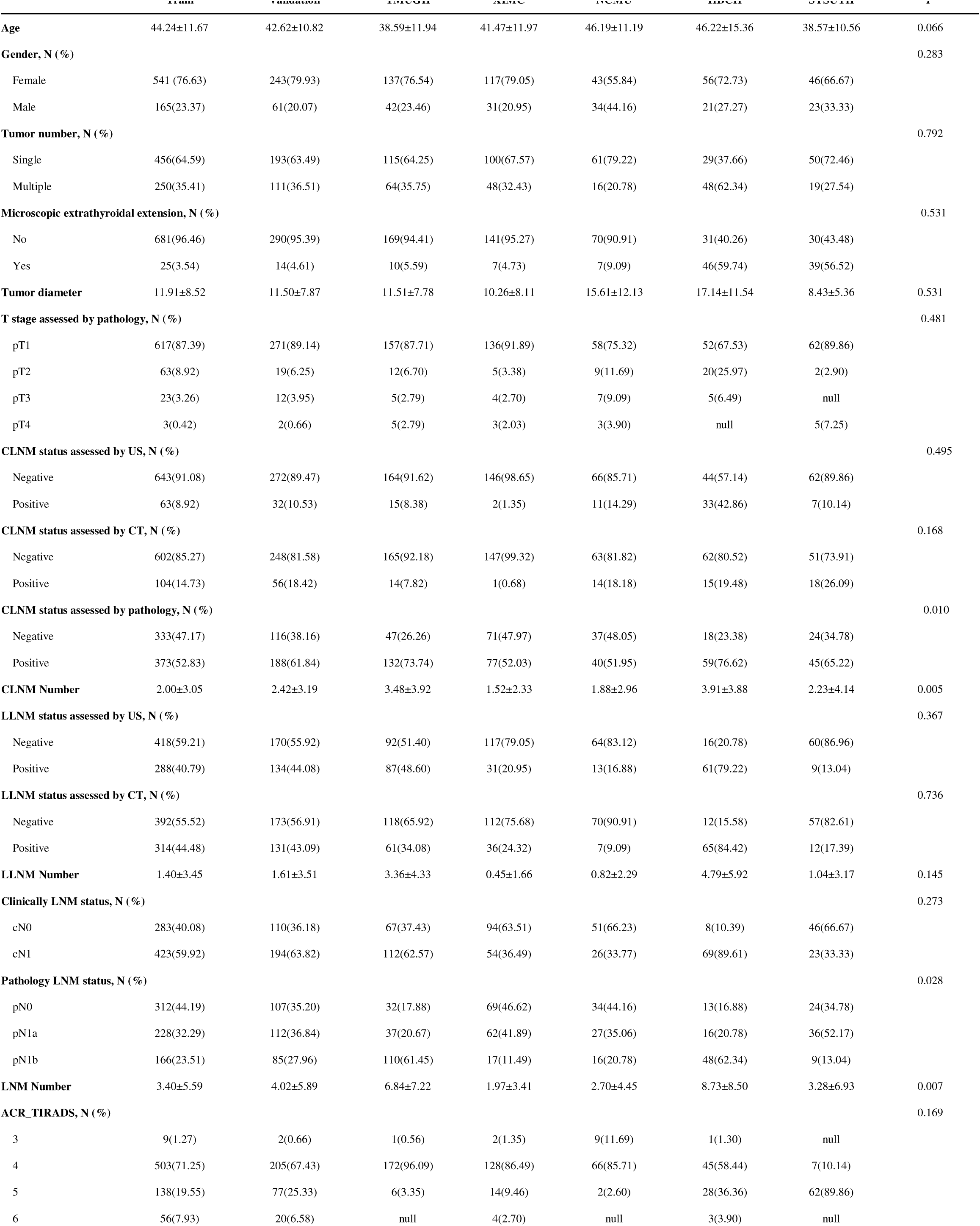
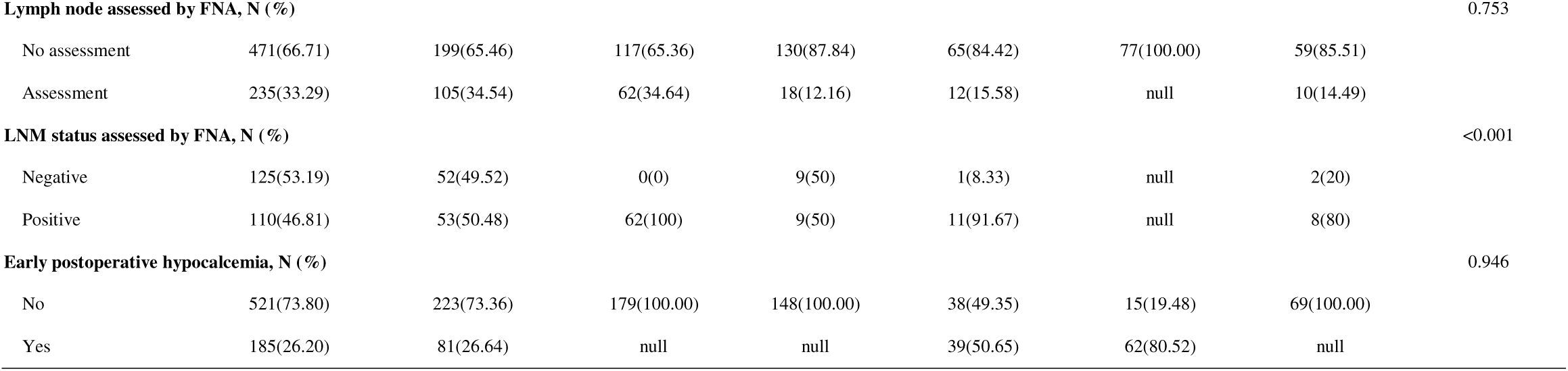
Baseline characteristics of the study cohorts.

### Slice Level Results

DenseNet201 demonstrated superior performance across the validation set compared to other CNN architectures (Supplementary Table 1). It achieved an AUC of 0.699 (95% CI: 0.6771-0.7206) on the validation cohort, surpassing ResNet18 (AUC: 0.648), ResNet50 (AUC: 0.642), ResNet101 (AUC: 0.690), and DenseNet121 (AUC: 0.695).

Additionally, DenseNet201’s specificity of 0.873 in the validation set highlights its effectiveness in reducing false positives compared to other architectures. To avoid information leakage from the test set to the model, we performed testing exclusively on the validation cohort.

Based on the observed results (Figure 2), DenseNet201 was selected as the base model for further comparisons involving Transformer-based feature fusion, ensemble learning, and MIL approaches. Its superior AUC across validation cohort underlines its robustness and suitability as a foundational architecture for subsequent model enhancements.

**Figure 2.**
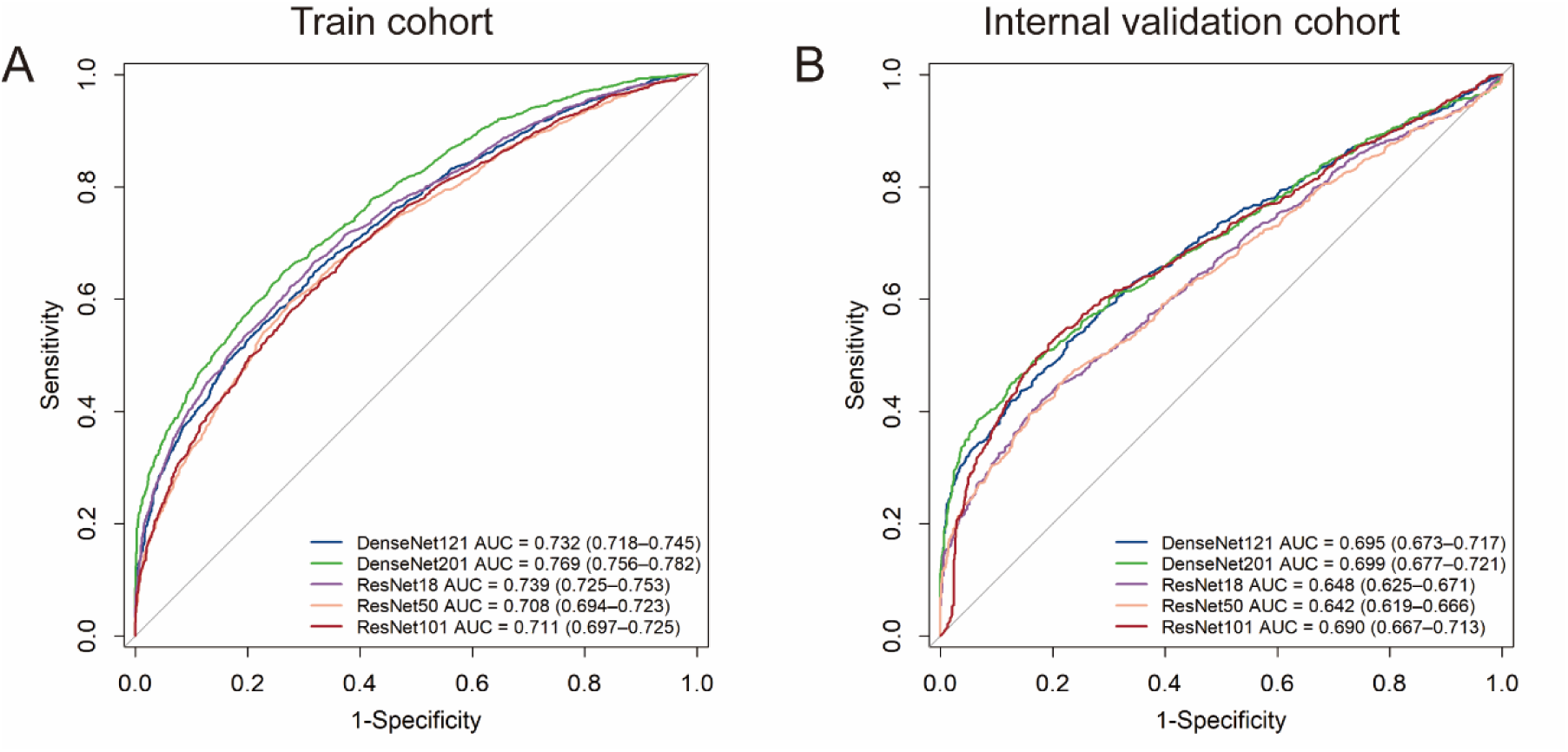
ROC curves of different CNN architectures for slice-level classification in the training and internal validation cohorts.

### Grad-CAM

To investigate the recognition capability of the deep learning model, gradient-weighted class activation mapping (Grad-CAM) was applied to visualize the activation maps of the final convolutional layer associated with class predictions (Figure 3). The highlighted regions indicate areas that most strongly influenced the model’s decision, thereby enhancing interpretability.

**Figure 3.**
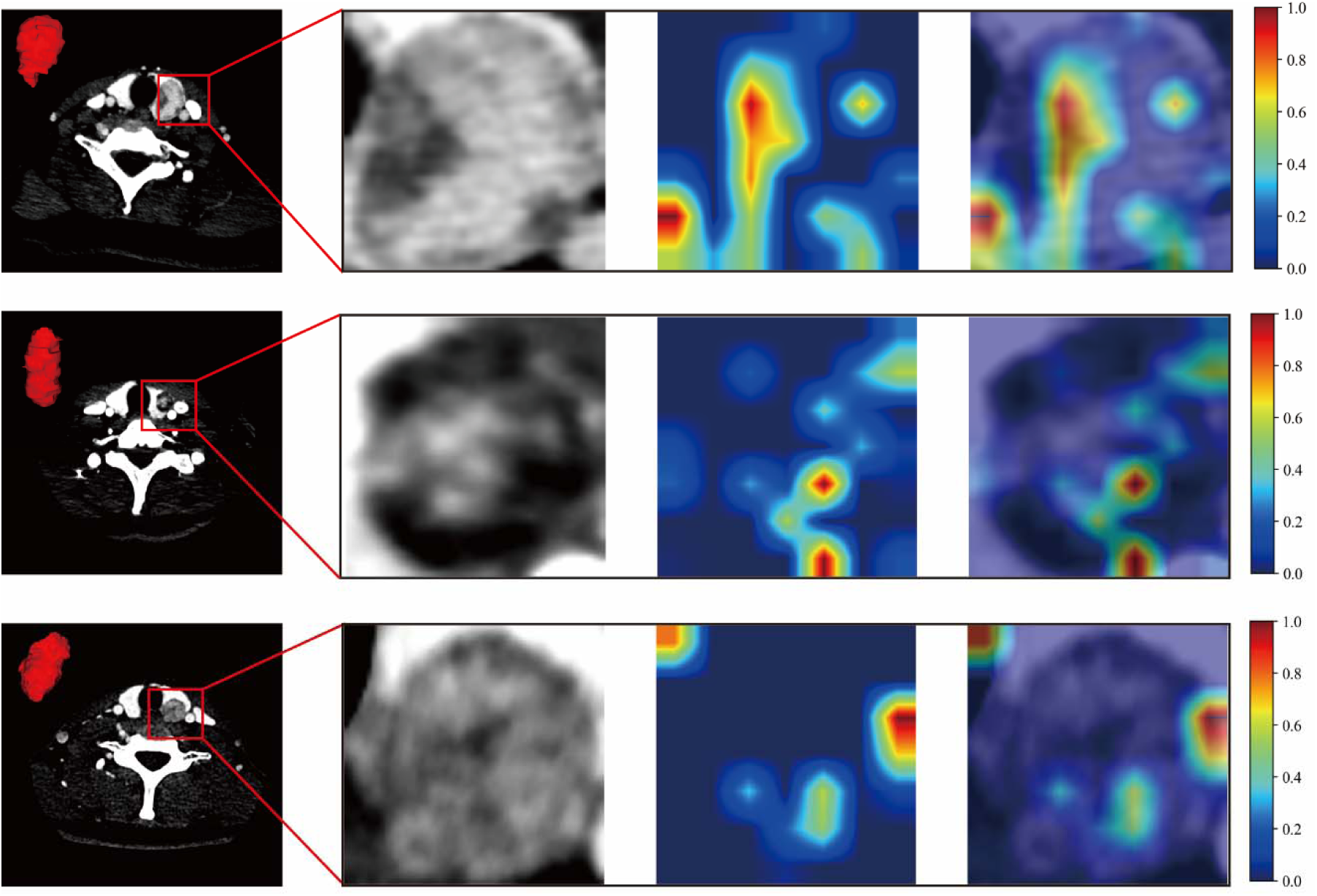
presents the Grad-CAM visualization of a sample, demonstrating that the attention map effectively focuses on the lesion area within the ROI.

### Transformer Based Patient Level Results

The Transformer model (named ThyLNT) consistently achieved higher AUC values compared to radiomics, ensemble, and MIL models across most cohorts (Table 2). For example, in the training cohort, ThyLNT attained an AUC of 0.882 (95% CI: 0.8577–0.9064), outperforming radiomics (0.802), ensemble (0.835), and MIL (0.863). Similarly, in the validation cohort, ThyLNT achieved an AUC of 0.787, exceeding MIL (0.715), ensemble (0.726), and radiomics (0.785). For the external test cohorts, AUC values ranged from 0.772 to 0.827, with the TMUGH cohort achieving the highest AUC of 0.827 (95% CI: 0.7643–0.8892), further demonstrating the model’s consistent performance across diverse datasets (Figure 4). Specificity was consistently high across cohorts, highlighting the model’s effectiveness in reducing false positives. For further details on the MIL and radiomics results, refer to Supplementary Materials 2B and 3A.

**Figure 4.**
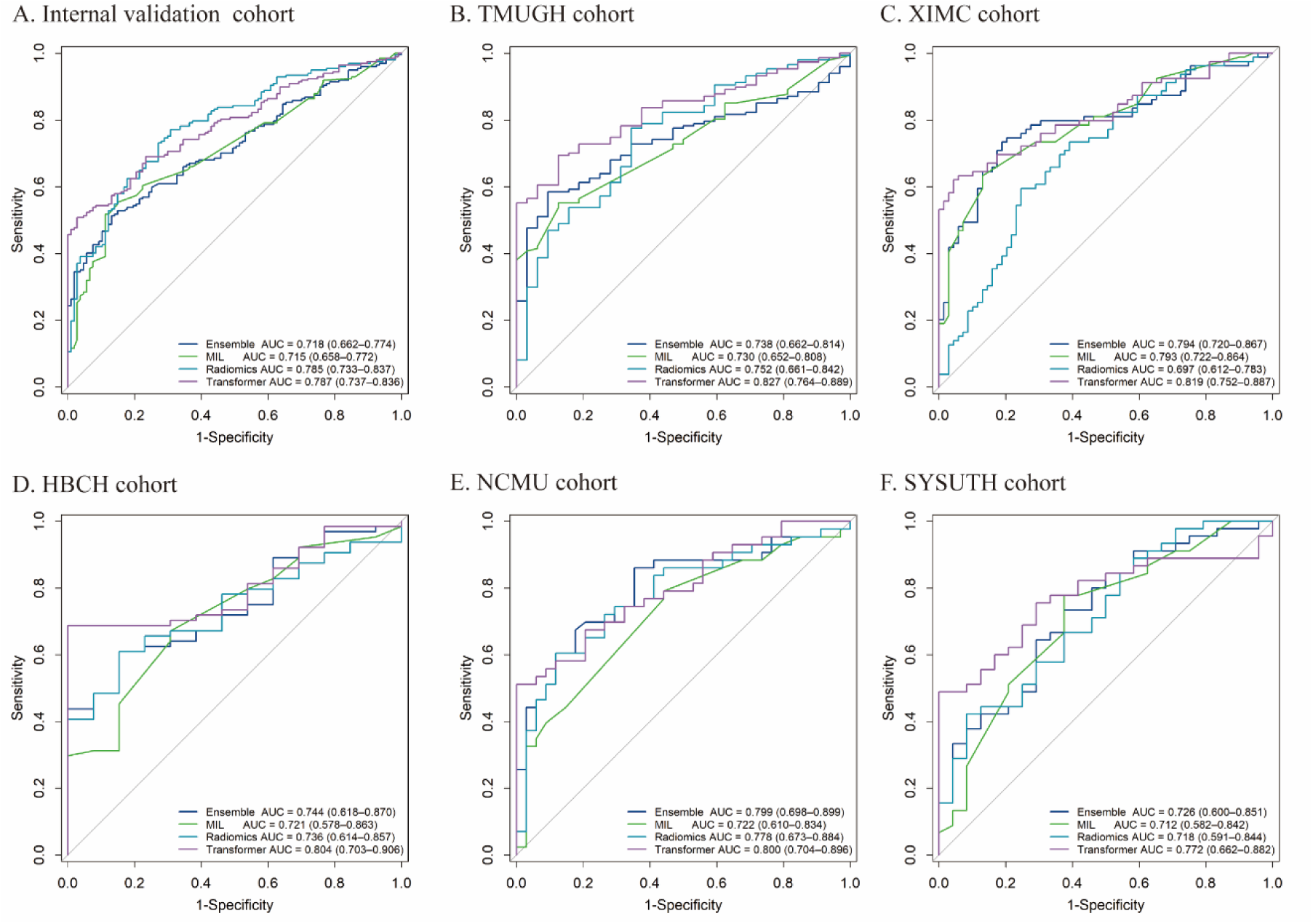
ROC curves of different models across the training, validation, and external cohorts.

**Table 2.**
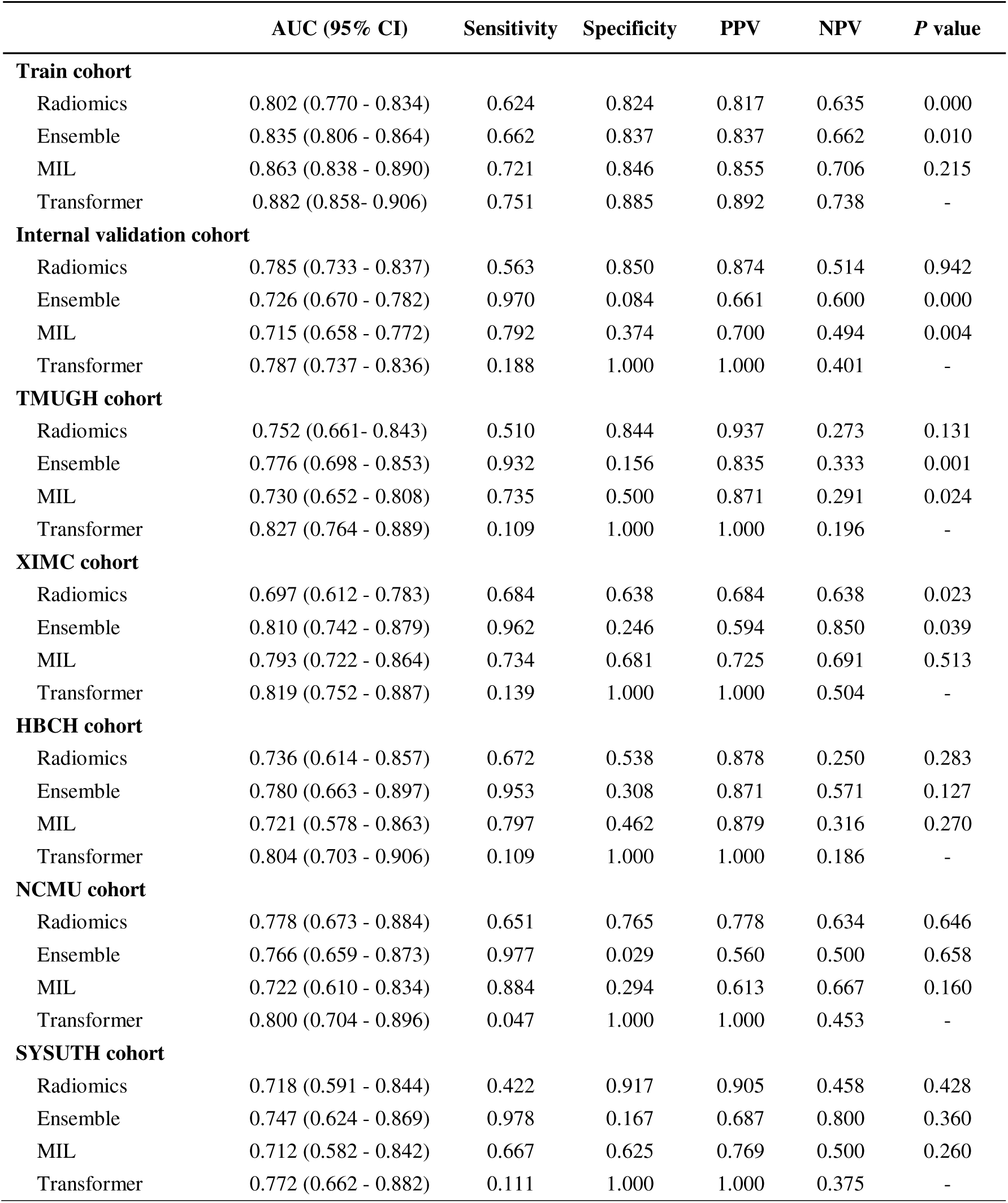
The performance metrics of different models across the training, validation, and external cohorts.

|  | AUC (95% CI) | Sensitivity | Specificity | PPV | NPV | P value |
| --- | --- | --- | --- | --- | --- | --- |
| **Train cohort** |  |  |  |  |  |  |
| Radiomics | 0.802 (0.770 - 0.834) | 0.624 | 0.824 | 0.817 | 0.635 | 0.000 |
| Ensemble | 0.835 (0.806 - 0.864) | 0.662 | 0.837 | 0.837 | 0.662 | 0.010 |
| MIL | 0.863 (0.838 - 0.890) | 0.721 | 0.846 | 0.855 | 0.706 | 0.215 |
| Transformer | 0.882 (0.858- 0.906) | 0.751 | 0.885 | 0.892 | 0.738 | - |
| **Internal validation cohort** |  |  |  |  |  |  |
| Radiomics | 0.785 (0.733 - 0.837) | 0.563 | 0.850 | 0.874 | 0.514 | 0.942 |
| Ensemble | 0.726 (0.670 - 0.782) | 0.970 | 0.084 | 0.661 | 0.600 | 0.000 |
| MIL | 0.715 (0.658 - 0.772) | 0.792 | 0.374 | 0.700 | 0.494 | 0.004 |
| Transformer | 0.787 (0.737 - 0.836) | 0.188 | 1.000 | 1.000 | 0.401 | - |
| **TMUGH cohort** |  |  |  |  |  |  |
| Radiomics | 0.752 (0.661- 0.843) | 0.510 | 0.844 | 0.937 | 0.273 | 0.131 |
| Ensemble | 0.776 (0.698 - 0.853) | 0.932 | 0.156 | 0.835 | 0.333 | 0.001 |
| MIL | 0.730 (0.652 - 0.808) | 0.735 | 0.500 | 0.871 | 0.291 | 0.024 |
| Transformer | 0.827 (0.764 - 0.889) | 0.109 | 1.000 | 1.000 | 0.196 | - |
| **XIMC cohort** |  |  |  |  |  |  |
| Radiomics | 0.697 (0.612 - 0.783) | 0.684 | 0.638 | 0.684 | 0.638 | 0.023 |
| Ensemble | 0.810 (0.742 - 0.879) | 0.962 | 0.246 | 0.594 | 0.850 | 0.039 |
| MIL | 0.793 (0.722 - 0.864) | 0.734 | 0.681 | 0.725 | 0.691 | 0.513 |
| Transformer | 0.819 (0.752 - 0.887) | 0.139 | 1.000 | 1.000 | 0.504 | - |
| **HBCH cohort** |  |  |  |  |  |  |
| Radiomics | 0.736 (0.614 - 0.857) | 0.672 | 0.538 | 0.878 | 0.250 | 0.283 |
| Ensemble | 0.780 (0.663 - 0.897) | 0.953 | 0.308 | 0.871 | 0.571 | 0.127 |
| MIL | 0.721 (0.578 - 0.863) | 0.797 | 0.462 | 0.879 | 0.316 | 0.270 |
| Transformer | 0.804 (0.703 - 0.906) | 0.109 | 1.000 | 1.000 | 0.186 | - |
| **NCMU cohort** |  |  |  |  |  |  |
| Radiomics | 0.778 (0.673 - 0.884) | 0.651 | 0.765 | 0.778 | 0.634 | 0.646 |
| Ensemble | 0.766 (0.659 - 0.873) | 0.977 | 0.029 | 0.560 | 0.500 | 0.658 |
| MIL | 0.722 (0.610 - 0.834) | 0.884 | 0.294 | 0.613 | 0.667 | 0.160 |
| Transformer | 0.800 (0.704 - 0.896) | 0.047 | 1.000 | 1.000 | 0.453 | - |
| **SYSUTH cohort** |  |  |  |  |  |  |
| Radiomics | 0.718 (0.591 - 0.844) | 0.422 | 0.917 | 0.905 | 0.458 | 0.428 |
| Ensemble | 0.747 (0.624 - 0.869) | 0.978 | 0.167 | 0.687 | 0.800 | 0.360 |
| MIL | 0.712 (0.582 - 0.842) | 0.667 | 0.625 | 0.769 | 0.500 | 0.260 |
| Transformer | 0.772 (0.662 - 0.882) | 0.111 | 1.000 | 1.000 | 0.375 | - |

DCA demonstrated that ThyLNT provided greater net benefit across a wide range of threshold probabilities in both the training and testing cohorts. In addition, PR curve analysis showed that ThyLNT achieved consistently higher AP values compared with the other models across most cohorts, indicating improved performance under class imbalance conditions. Calibration analysis further confirmed good agreement between predicted probabilities and observed outcomes, and the Hosmer–Lemeshow test did not suggest significant miscalibration (P = 0.757 in the training cohort, P = 0.096 in the validation cohort, and P = 0.349–0.728 in the external cohorts). Furthermore, pairwise comparisons using the DeLong test indicated that ThyLNT showed statistically significant improvements over certain comparator models across multiple cohorts (Supplementary Figure 5).

We further compared the performance of ThyLNT as an intraoperative decision-support tool with conventional preoperative thyroid imaging modalities, namely neck US and CT. In the validation cohort, ThyLNT achieved a significantly higher AUC than US alone, CT alone, and their combination (all P < 0.001). In the external test cohorts, ThyLNT consistently outperformed at least one of the imaging modalities (Figure 5A). Notably, integrating ThyLNT with US and CT did not yield additional performance gains (Supplementary Table 5), likely because ThyLNT already captured information highly relevant to the outcome, leaving limited marginal contribution from imaging variables.

**Figure 5.**
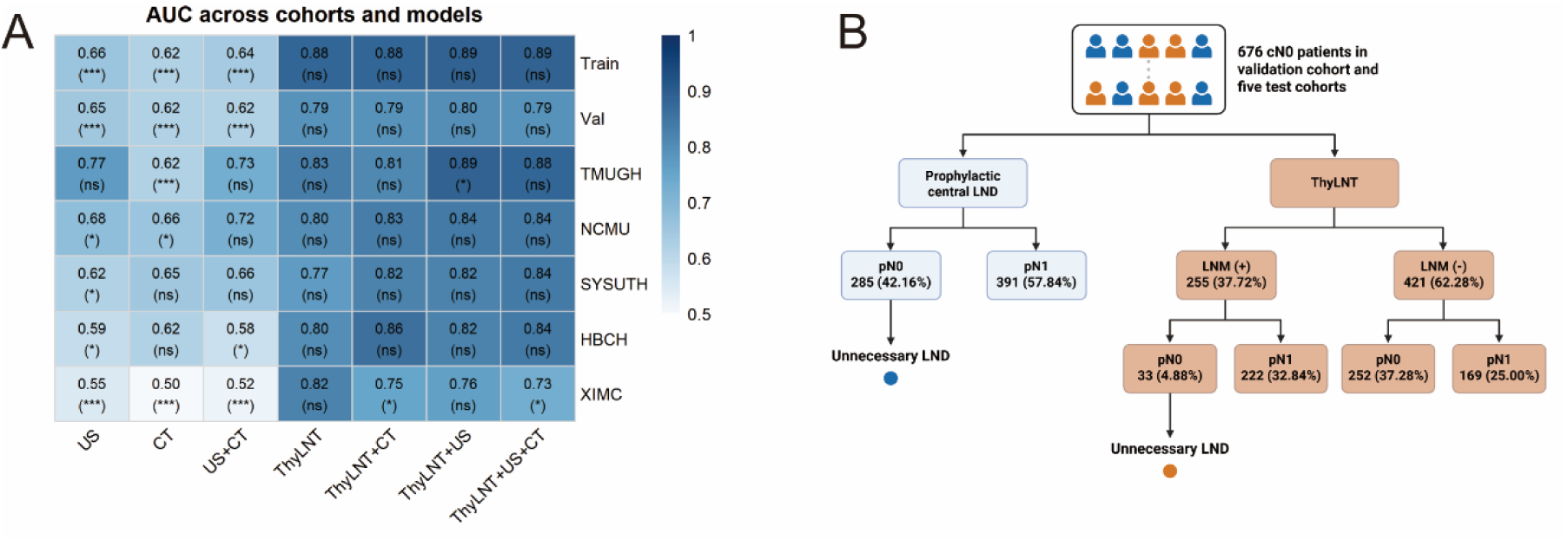
(A) AUC heatmap across all cohorts. Colors indicate AUC (color bar). Each cell reports AUC followed by the DeLong test P value in parentheses (vs ThyLNT). Significance: *P < 0.05, **P < 0.01, ***P < 0.001; ns. (B) Recommendation for LND in patients with cN0 PTC based on the ThyLNT model.

Across the validation cohort and five test cohorts, 676 cN0 patients underwent prophylactic central LND. When LND decisions were simulated based on ThyLNT predictions, the rate of unnecessary LND decreased substantially from 52.16% to 4.88% (Figure 5B), suggesting that ThyLNT may help reduce potential overtreatment and the risk of procedure-related morbidity.

To explore the molecular basis underlying the model predictions, we performed differential expression and enrichment analyses using both the model-predicted groups and the true LNM-based clinical groups. Notably, although the grouping was defined by LNM status, the transcriptomic analysis was performed using primary tumor lesions (the ThyLNT model also extracts features from the primary-tumor ROI); as reported previously in this setting, no DEGs were identified using adjusted P values^13^, we adopted a less stringent criterion using nominal P values (P < 0.05) to identify differentially expressed genes (DEGs), consistent with previous reports. In the model-predicted grouping, a total of 931 upregulated genes and 52 downregulated genes were identified (Figure 6A), whereas in the true clinical grouping, 900 upregulated genes and 78 downregulated genes were identified (Supplementary Figure 6A). GO and KEGG enrichment analyses showed that DEGs from both groupings were consistently enriched in cell motility–related pathways and associated biological processes (Figure 6C and Supplementary Figure 6B), indicating that these metastasis-associated programs were effectively captured by the model.

**Figure 6.**
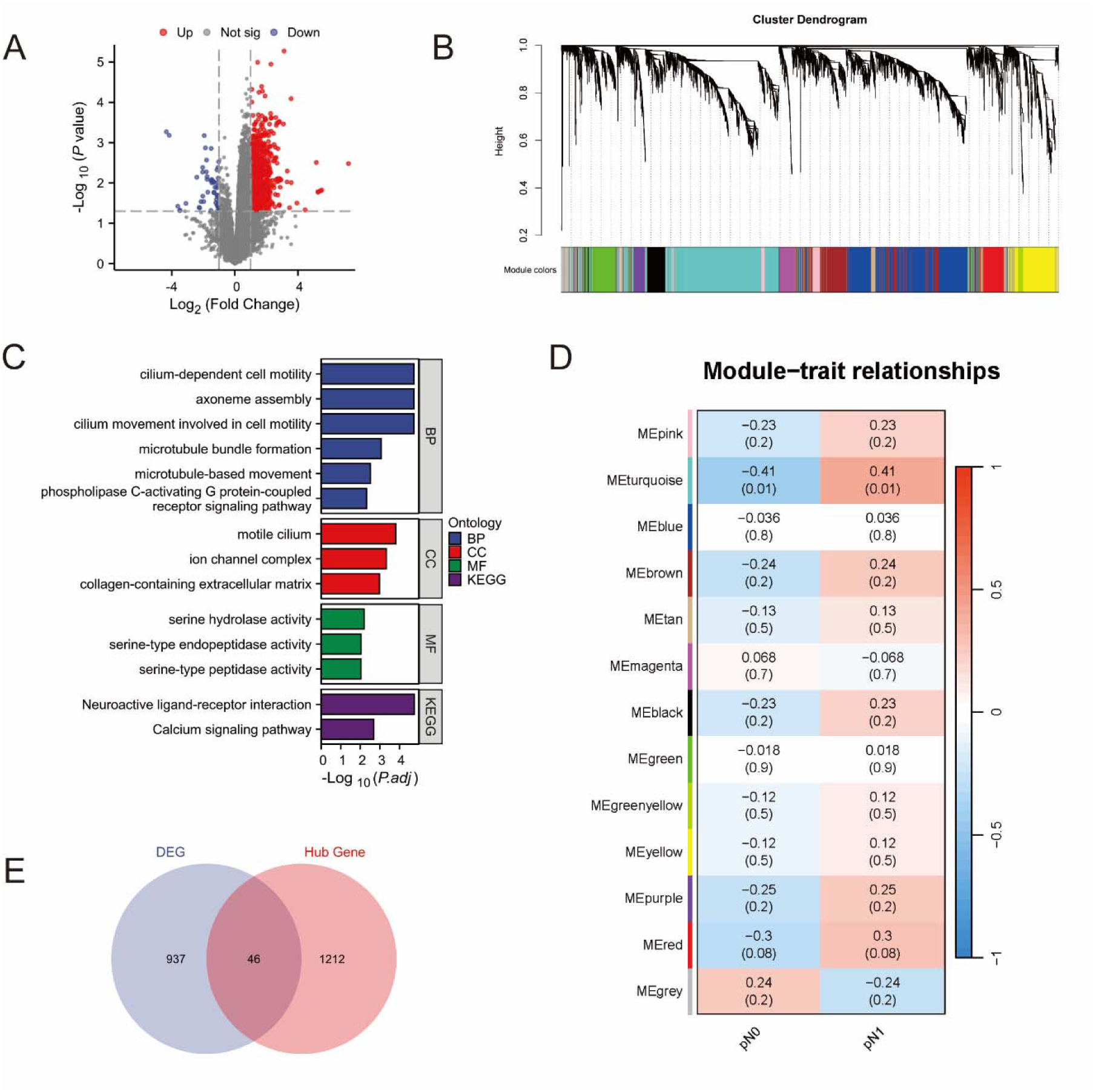
(A) Volcano plot of DEGs between predicted groups. Red dots indicate upregulated genes, blue dots indicate downregulated genes, and gray dots represent non-significant genes. (B) Hierarchical clustering dendrogram of genes generated by WGCNA. Genes with similar expression patterns were grouped into distinct modules, indicated by different colors. (C) GO and KEGG enrichment analysis of DEGs. Biological Process (BP), Cellular Component (CC), Molecular Function (MF), and KEGG pathway categories are shown. (D) Module-trait relationship heatmap showing correlations between module eigengenes and LNM status (pN0 vs pN1). Correlation coefficients and corresponding P values are indicated in each cell. (E) Venn diagram showing the overlap between DEGs and hub genes identified from WGCNA modules.

Considering the potential cooperative effects among genes, WGCNA was performed to identify gene co-expression modules (Figure 6B). Module eigengenes were correlated with model prediction scores and LNM status to identify relevant modules (Figure 6D). To further refine candidate genes, we intersected the DEGs with genes from the significantly associated modules (Figure 6E). The overlapping genes were subsequently ranked according to their correlation coefficients with the model prediction scores.

Among these, VEGFA exhibited the strongest correlation with the model predictions, highlighting it as a key candidate gene associated with the model-derived risk stratification.

Single-cell RNA sequencing analysis identified distinct cellular populations within the tumor microenvironment (Figure 7A). Pseudotime analysis revealed branched developmental trajectories, with VEGFA significantly upregulated along specific branches in lymph node metastatic cells (Figure 7B), indicating its association with metastasis-related state transitions. Spatial transcriptomic analysis in locally advanced PTC patients further demonstrated that VEGFA exhibited focal high expression within the tissue, predominantly enriched in thyrocyte-dominated regions and spatially adjacent to myeloid cell– and fibroblast-enriched areas (Figure 7C), supporting its involvement in tumor microenvironment remodeling. Cell-cell communication analysis showed a global intercellular interaction network (Figure 7D), and VEGF pathway-specific role analysis indicated that VEGF signaling was mainly initiated by Thyrocytes, while Endothelial cells served as the principal signal receivers (Figure 7E), consistent with the canonical VEGF-VEGFR axis. Myeloid and Fibroblast populations functioned as mediators within the network. Ligand-receptor contribution analysis identified VEGFB-VEGFR1 as the dominant signaling axis, followed by VEGFA-associated interactions. Furthermore, in silico knockout of VEGFA using scTenifoldKnk revealed significant network perturbations, with affected genes enriched in angiogenesis and epithelial–mesenchymal transition (EMT) pathways (Figure 7F).

**Figure 7.**
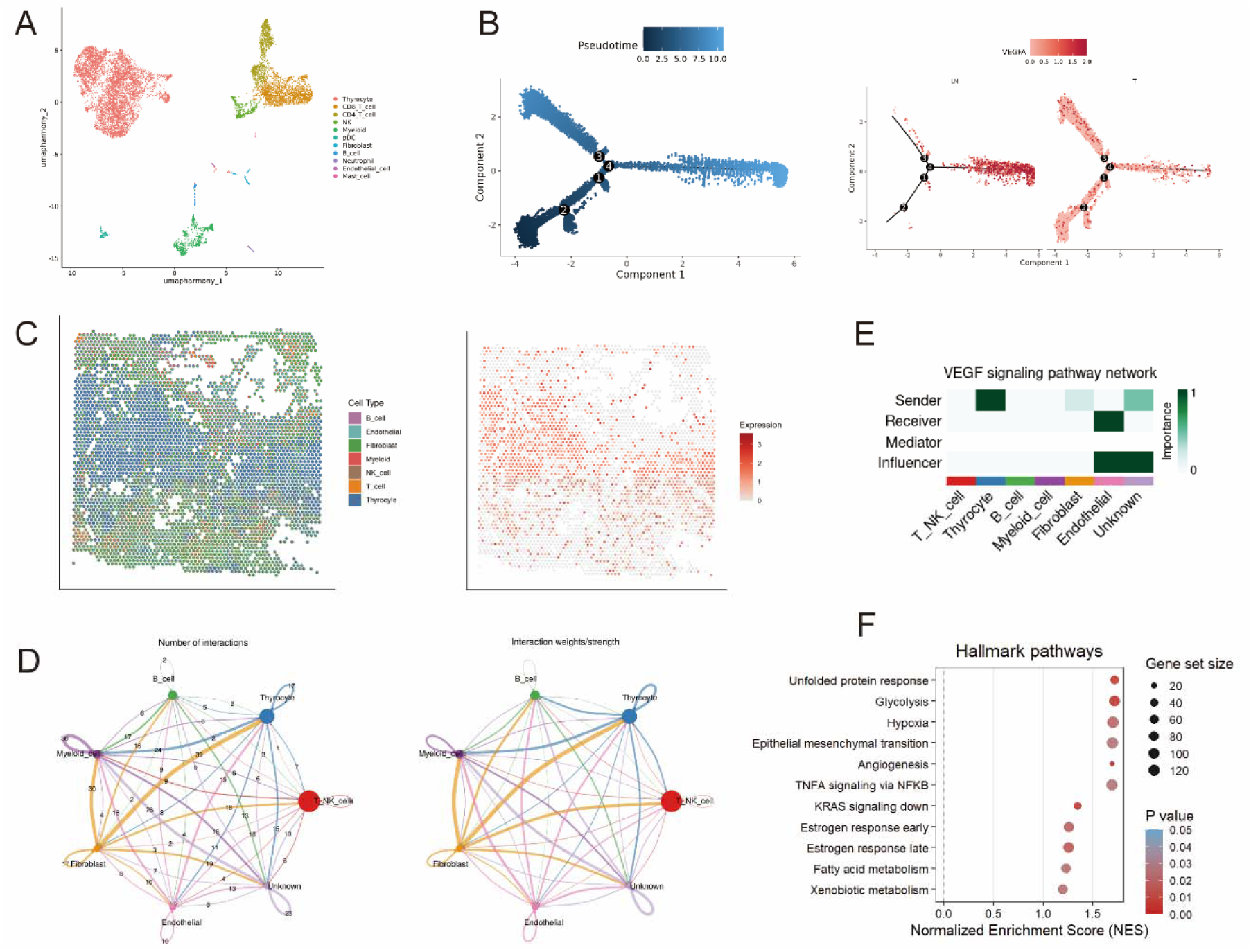
(A) UMAP visualization of major cell populations identified from integrated single-cell RNA-seq data. (B) Pseudotime trajectory analysis and VEGFA expression dynamics in LNM and primary tumor (T) cells. (C) Spatial transcriptomics showing cell type distribution and spatial expression pattern of VEGFA. (D) Global cell-cell communication network illustrating overall interaction number and strength among major cell types. (E) Functional role analysis of the VEGF signaling pathway indicating the relative importance of each cell type as sender, receiver, mediator, and influencer. (F) Functional enrichment analysis of genes perturbed by in silico knockout of VEGFA using scTenifoldKnk.

Spatial metabolomics analysis revealed marked metabolic heterogeneity between LNM and non-LNM tissues. Metabolites with high VIP scores derived from the OPLS-DA model exhibited distinct spatial distribution patterns, indicating localized metabolic remodeling within metastatic tissues (Figure 8A). Differential analysis identified 95 upregulated and 14 downregulated metabolites in LNM samples compared with non-LNM tissues (Figure 8B), suggesting extensive metabolic reprogramming associated with metastatic progression. Hierarchical clustering further demonstrated clear separation between LNM and non-LNM samples based on metabolite abundance profiles (Figure 8C). The altered metabolites were primarily enriched in amino acids and their derivatives, organic acids, fatty acids, glycerolipids, and carbohydrate-related metabolites. KEGG pathway enrichment analysis revealed involvement of lipid-related metabolic pathways, including glycerophospholipid metabolism, arachidonic acid metabolism, linoleic acid metabolism, and choline metabolism in cancer (Figure 8D), highlighting lipid metabolic dysregulation as a key feature of metastatic tissues. Correlation network analysis demonstrated extensive and coordinated interactions among differential metabolites (Figure 8E), suggesting that metastatic progression is characterized by systematic metabolic network remodeling rather than isolated metabolic alterations.

**Figure 8.**
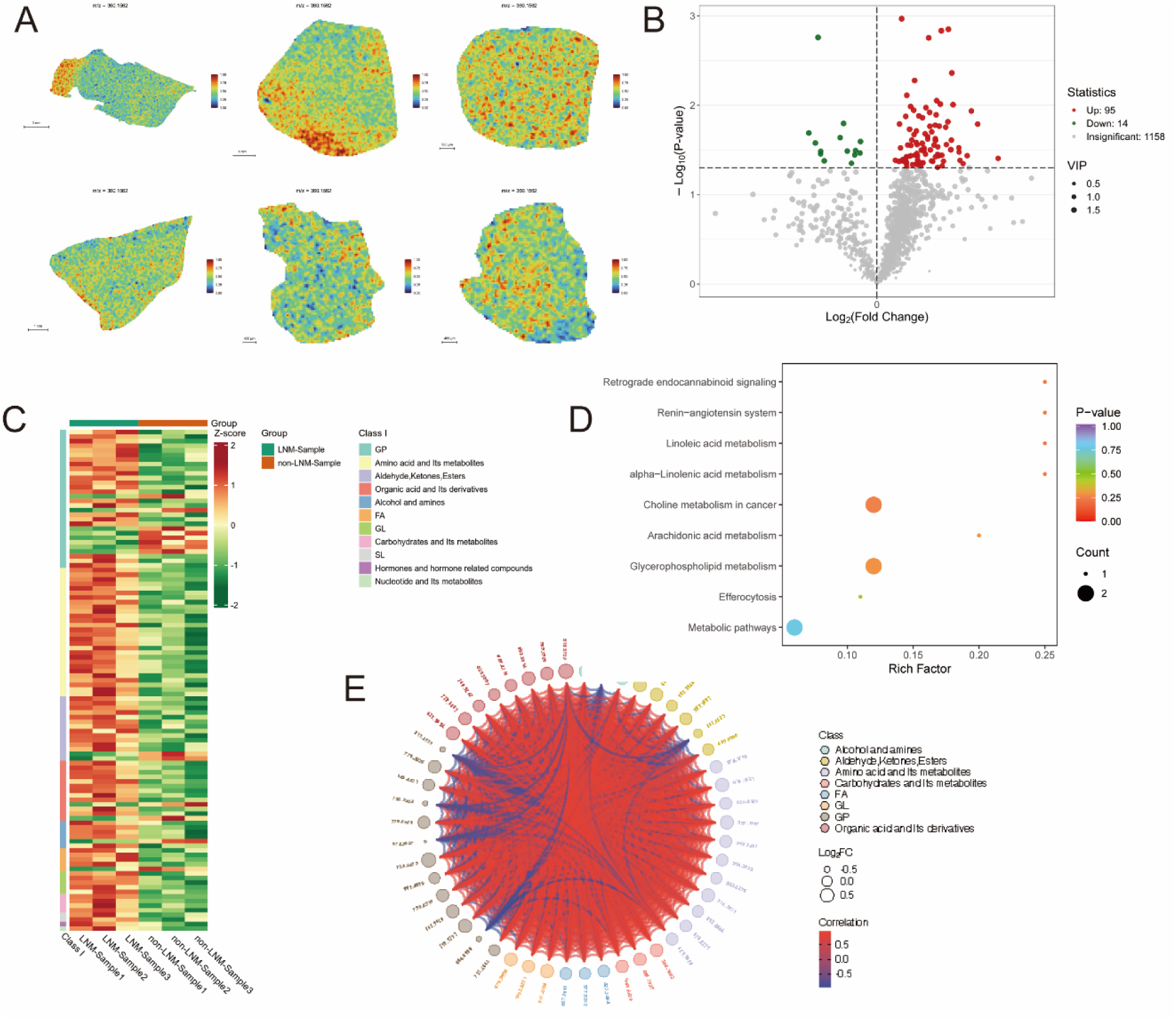
(A) Spatial distribution maps of representative metabolites across tissue sections from LNM and non-LNM samples. Color scale indicates relative metabolite abundance. (B) Volcano plot showing differentially expressed metabolites between LNM and non-LNM groups. Red dots represent significantly upregulated metabolites, green dots represent downregulated metabolites, and gray dots indicate non-significant features. (C) Heatmap of differentially abundant metabolites across samples, grouped by metabolic class. Z-scores represent normalized metabolite abundance. (D) KEGG pathway enrichment analysis of differential metabolites. Bubble size indicates metabolite count, and color represents adjusted p-value. (E) Correlation network of differential metabolites. Node size represents fold change, color indicates metabolic class, and edge color represents correlation coefficient.

## Discussion

Accurate preoperative prediction of cervical LNM in PTC remains a central challenge influencing decisions regarding the extent of surgery. Although neck US and CT have been widely used for preoperative staging, real-world multicenter data indicate that both imaging modalities have inherent structural limitations in detecting cervical metastases. Central compartment lymph nodes are affected by anatomical obscuration, small size, and interference from adjacent tracheal and osseous structures, resulting in limited preoperative sensitivity and a relatively high rate of occult metastasis^14^. In contrast, although lateral lymph nodes are relatively more accessible, missed diagnoses or misinterpretations may still occur in cases of micrometastasis, atypical morphology, or inflammatory reactive enlargement^15^. Therefore, reliance solely on preoperative imaging is insufficient to fully and accurately determine cervical lymph node status. In this context, international guidelines (e.g., the 2015 ATA guidelines) do not recommend routine prophylactic central neck dissection (pCND) for clinically cN0 PTC, emphasizing a risk-stratified and individualized approach that balances potential benefits against surgery-related complications^5^. However, in China and certain other Asian regions, due to the relatively high rate of occult metastasis and limitations in preoperative assessment,

LND_—_particularly central compartment dissection_—_remains more aggressive in clinical practice. Relevant meta-analyses further suggest that conclusions regarding the risk_–_benefit balance vary across regions^16^, resulting in substantial geographic differences in treatment strategies and ongoing controversy.

ThyLNT extracts deep features based on 2.5D slice sampling from CT images and integrates multi-slice information through feature-level Transformer fusion, maintaining stable discriminative performance in multicenter external validation and overall outperforming ensemble learning and MIL approaches. In simulations involving the cN0 population, it also significantly reduced the proportion of unnecessary LND. These findings suggest that the model has the potential to provide a quantifiable preoperative risk stratification tool for determining whether LND is necessary in cN0 patients. It should be noted that this study used overall LNM as the endpoint without distinguishing between central and lateral compartment metastasis; therefore, when applying the overall prediction results to interpret decisions regarding central neck dissection, the conclusions may theoretically be influenced by skip metastasis (negative CLNM with positive LLNM). However, previous studies have shown that the incidence of skip metastasis is relatively low^17^, and its impact on the overall simulation conclusions is expected to be limited.

The 2.5D strategy achieves a balance between contextual representation and computational cost; however, imaging signals associated with metastasis are often heterogeneously distributed across slices: Certain slices may reflect capsular invasion, disruption of the tumor_–_capsule interface, or alterations in the surrounding interstitial tissue, whereas others may contain limited informative content. Within this framework of patient-level supervision and slice-level representation learning, improvements in predictive performance depend on the effective integration of cross-slice information rather than the discriminative power of any single slice. MIL methods based on pooling or attention pooling typically assume a degree of interchangeability among instances.

However, in CT axial sequences, slices are not fully interchangeable, as their spatial continuity and structural variations across adjacent levels may themselves carry critical evidence. The self-attention mechanism of the Transformer explicitly models dependencies and mutual reinforcement among slice tokens, a concept consistent with the advantages of Vision Transformers in capturing long-range dependencies and multi-view fusion^10^, and which has been repeatedly shown in medical imaging to outperform traditional convolutional architectures in cross-regional information integration^18, 19^.

Therefore, the observed pattern in this study_—_moderate slice-level performance but significantly improved patient-level performance after Transformer fusion_—_more likely reflects that lymph node metastatic risk represents a composite imaging phenotype distributed across slices, rather than a localized event determined by a single most discriminative slice.

At the level of clinical translation, this study demonstrated that integrating ThyLNT with conventional imaging assessments (US/CT reports) did not confer further performance gains. This may be attributed to the fact that ThyLNT has already incorporated the majority of outcome-relevant imaging information. Consistent with previous evidence, imaging phenotypes of the primary lesion alone are sufficient to effectively predict lymph node metastasis^20^. Moreover, given that the risk_–_benefit balance of pCND may vary across regions, and that meta-analyses have reported inconsistent conclusions regarding efficacy, recurrence, and complications^21^, employing ThyLNT as a risk stratification tool with center-specific threshold optimization based on tolerance for missed diagnosis versus overtreatment may be more aligned with real-world clinical pathways than adopting a single fixed threshold.

Previous studies suggest that LNM in PTC is not merely a consequence of anatomical spread but is closely associated with tumor microenvironment remodeling, including activation of lymphangiogenic and angiogenic signaling pathways^22^, enhanced cellular migration and invasion, and alterations in immune cell infiltration and intercellular communication^23^. In addition, tumor metabolic reprogramming, particularly abnormalities in lipid metabolism, has been implicated in metastatic formation and the establishment of the metastatic niche^24^. These molecular and microenvironmental alterations may leave learnable microscopic phenotypic signatures on CT imaging_—_such as changes in tumor margins, interstitial architecture, and tissue texture_—_and imaging features may also reflect underlying gene expression patterns and tumor molecular pathway states^25^. The association between VEGF-related signaling and metastatic risk has been supported in previous PTC literature, with systematic reviews and meta-analyses reporting correlations between VEGF expression and LNM^22^. In this study, network analysis of bulk RNA-seq data, along with single-cell and spatial transcriptomics, further aligned the VEGF axis with cellular communication architecture, thereby enhancing the biological coherence of the “imaging phenotype-pathway-microenvironment remodeling” framework. Furthermore, spatial metabolomics revealed significant alterations and coordinated network activity in lipid metabolism-related pathways, which, together with angiogenic and lymphangiogenic signaling, constitute a candidate mechanistic framework for the metastatic microenvironment and provide direction for future histological and functional validation.

This study also has several limitations that warrant clarification. The predictive endpoint did not distinguish between CLNM and LLNM but modeled overall LNM as a composite outcome. However, from a surgical perspective, the indications, imaging detectability, and potential benefits differ between these compartments. Recent reviews have also emphasized regional involvement patterns from the central to lateral compartments and phenomena such as skip metastasis in PTC, suggesting that compartment-specific modeling may offer additional value^23^. Therefore, future studies with larger sample sizes incorporating compartment-specific or multi-task models for CLNM and LLNM may enhance the model’s capacity to directly inform decisions regarding the extent of surgery, such as central versus lateral neck dissection.

## Data Availability

The data supporting the conclusions of this study can be found within the article and its Supplementary Information files, and are also available from the corresponding author upon reasonable request.

## Acknowledgments

Funding

This work was supported by the National Natural Science Foundation of China (No. 82203392), the Natural Science Foundation of Hubei Province (JCZRMS202602288), and the Medical Artificial Intelligence General Program of Tongji Hospital 2025 (No. AI2025A02).

## Author contributions

Shaojie Xu, Xingmin Yan, Yuhang Su and Jia Qi are co-first authors who contributed equally to this work. Shaojie Xu was responsible for the conceptualization of the study, development of methodology, formal analysis and writing of the original draft; Xingmin Yan and Yuhang Su participated in the development of methodology, formal analysis and writing of the original draft; Jia Qi provided research resources and engaged in data curation. Zhihao Wei and YISIKANDAER YALIKUN also undertook the work of providing research resources and data curation. Xingyu Chen, Yulin Li, Hao Xiong and Jiamei Jiang completed the visualization work of this study through ROI drawing. Ziying Chen, Hanning Li and Xingyin Li assisted with formal analysis and the review & editing of the manuscript. Yiqing Xi, Wei Li, Xingrui Li and Yaying Du are co-corresponding authors who contributed equally in this work, all of whom provided research resources, conducted data curation, supervised the entire study, and offered critical revisions to the manuscript.

## Ethics declarations

### Ethics approval and consent to participate

The study was approved by the ethics committee of Tongji Hospital, Tongji Medical College, Huazhong University of Science and Technology (No. TJ-IRB202404008), with the requirement for informed consent being waived.

## Competing interests

The authors declare no competing interests.

## Notes

### Competing Interest Statement

The authors have declared no competing interest.

