## Supplementary material for "A Transformer-Based 2.5D Deep Learning Model for Preoperative Prediction of Lymph Node Metastasis in Papillary Thyroid Carcinoma": supplemmentary materials

### 1A. Details of Deep Learning Training Process

**Data Augmentation**: In this study, we applied Z-score normalization to standardize the intensity distribution across RGB channels in images, which were then used as input for our model. During the training phase, we employed real-time data augmentation techniques, such as random cropping and horizontal and vertical flipping. For the test images, we restricted the preprocessing to normalization only, without applying any augmentation.

**Data Normalization**: We normalized the grayscale values of the image slices using a min-max transformation to adjust the range to [-1, 1]. Each cropped subregion image was resized to 224 × 224 pixels using nearest neighbor interpolation, ensuring compatibility with the input requirements of our deep learning models.

**Training Parameters**: To optimize the learning process for our specific image dataset, we adjusted the learning rate using a cosine decay strategy, detailed in the following equation:

$$\eta_{t}=\eta_{min}^{i}+\frac{1}{2}\left( \eta_{max}^{i}-\eta_{min}^{i} \right)\left( 1+cos\left( \frac{T_{cur}}{T_{i}}\pi\right) \right)$$

where $\eta_{min}^{i}$ is set to 0, and $\eta_{max}^{i}$ to 0.01, with $T_{i}$ representing the number of iteration epochs. We utilized SGD (Stochastic Gradient Descent) as the optimizer, and softmax cross entropy as the loss function.

### 2A. Details of Transformer Fusion

The process of using a Transformer for feature fusion in treatment efficacy evaluation can be summarized in the following steps:

1. **Feature Extraction**: Extract deep learning features from seven distinct cross-sectional images, namely the maximum cross-section, +1, +2, +4, -1, -2, and -4 sections.
2. **Tokenization**: Treat each set of extracted features as a token, analogous to a word in a sentence.
3. **Embedding**: Pass the feature tokens through an embedding layer to project them into a higher-dimensional space, facilitating more effective feature interaction.
4. **Transformer Encoding**:
   - **a. Self-Attention**: Apply a multi-head self-attention mechanism to weigh the importance of different features dynamically, enabling the model to focus on the most relevant information.
   - **b. Feed-Forward Network**: Utilize a position-wise feed-forward network to further refine the feature representation.
5. **Feature Fusion**: The output of the Transformer encoder is a set of fused features, which encapsulate the combined information from all seven cross-sectional images.
6. **Prediction**: Pass the fused features through a final classification or regression layer, depending on the specific task, to make the final prediction regarding treatment efficacy.

### 2B. Details Multi-Instance Learning-Based Feature Fusion

In our study, we implemented two multi-instance learning fusion techniques. Using 2.5D deep learning models, we created Predict Likelihood Histograms (PLH) that map out the predictive probabilities and labels for each slice, offering a probabilistic summary of the prediction landscape. We also applied a Bag of Words (BoW) approach, segmenting each image into slices and extracting data to compile seven predictive results per sample, using the Term Frequency-Inverse Document Frequency (TF-IDF) method for analysis. Additionally, we enhanced our model by integrating PLH and BoW features with radiomic data, leveraging diverse data sources to improve the representational power and accuracy of our classification tasks.

Our multi-instance learning approach aimed to enhance predictive accuracy by integrating various data points from a single sample into a comprehensive feature set, involving:

1. **Slice Prediction**: Each slice was analyzed using the deep learning model to derive probabilities and labels, denoted as $Slice_{prob}$ and $Slice_{pred}$, retained to two decimal places.
2. **Multi Instance Learning Feature Aggregation**:

**Histogram Feature Aggregation**:

- Distinct numbers were treated as "bins" to count occurrences across types.
- Frequencies of $Slice_{prob}$ and $Slice_{pred}$ in each bin were tallied and normalized using min-max normalization, resulting in $Histo_{prob}$ and $Histo_{pred}$.

**Bag of Words (BoW) Feature Aggregation**:

- A dictionary was constructed from unique elements in $Slice_{prob}$ and $Slice_{pred}$.
- Each slice was represented as a vector noting the frequency of each dictionary element, with a TF-IDF transformation applied to emphasize informative features.
- This resulted in a BoW feature representation for each slice, encapsulating both the presence and significance of features.

3. **Feature Early Fusion**: We integrated $Histo_{prob}$, $Histo_{pred}$, $Bow_{prob}$, and $Bow_{pred}$ using a feature concatenation method ($\oplus$), combining these into a single comprehensive feature vector:

$$feature_{fusion}=Histo_{prob}\oplus Histo_{pred}\oplus Bow_{prob}\oplus Bow_{pred}$$

For the aggregated multi-instance learning features, we utilized dimensionality reduction techniques such as t-tests, correlation coefficients, and Least Absolute Shrinkage and Selection Operator (LASSO) regularization to refine our feature set (Supplementary Figure 3). These features were modeled using popular machine learning algorithms including Logistic Regression and ET (Supplementary Table 2). To address sample imbalance, we employed the SMOTE method during the training process. To ensure model robustness, we applied 5-fold cross-validation within the training dataset and optimized hyperparameters via Grid-Search.

### 2C. Details Different Ensemble Fusion Method

Leveraging the concept of ensemble methods, we combined the predicted probabilities from different cross-sectional images of the same patient to enhance predictive performance and robustness. Specifically, we implemented two distinct approaches to perform this ensemble fusion: using the maximum and the average values of the predictions.

The first approach involves taking the maximum predicted probability among all slices for a given patient. This method tends to highlight the most abnormal features detected across slices, which might indicate the presence of pathological findings more assertively. The mathematical representation for the maximum ensemble method is as follows:

$$P_{\text{max}}=max\left( P_{+1},P_{+2},P_{+4},P_{max\_roi},P_{-1},P_{-2},P_{-4} \right)$$

The second approach averages the predicted probabilities across all slices. This method provides a more balanced view that incorporates contributions from all slices, potentially smoothing out any anomalies that might appear in a single slice. The formula for calculating the average predicted probability is:

$$P_{\text{mean}}=\frac{1}{n}\sum\left( P_{+1},P_{+2},P_{+4},P_{max\_roi},P_{-1},P_{-2},P_{-4} \right)$$

where $P_{\text{max}}$ is the maximum probability , where$P_{\text{mean}}$ represents the average probability and$P_{+1},P_{+2},P_{+4},P_{max\_roi},P_{-1},P_{-2},P_{-4}$are the predicted probabilities for each slice.

Detailed comparisons between these two ensemble methods are available, offering insights into their respective advantages and limitations in the context of medical image analysis (Supplementary Table 3). These comparative analyses help in selecting the most appropriate ensemble strategy based on specific clinical and diagnostic requirements.

### 3A. Results of Radiomics Signature

**Feature extraction**

In our analysis, we extracted omics features from CT imags and categorized them into three main types: (I) Geometry, reflecting tumor shape; (II) Intensity, based on the first-order statistical distribution of voxel intensities; and (III) Texture, describing spatial intensity patterns. Image transformations, such as Laplacian of Gaussian (LoG) and Wavelet, were applied to enhance feature representation. For texture features, we used methods like GLCM, GLRLM, GLSZM, and NGTDM (Supplementary Figure 4A). All features were extracted with pyradiomics (v3.0.1) following IBSI standards.

Feature fusion was applied to the multi-modal features, integrating data from various imaging modalities to generate a more holistic and enriched feature set. This fusion process allowed for the combination of complementary information from each modality, capturing distinct tumor characteristics that may not be fully represented by individual modalities alone.

**Feature selection**

In our methodology, we first standardized all extracted features using Z-score normalization to ensure a consistent scale for analysis. A t-test was then applied to these normalized features, retaining those with a p-value below 0.05, indicating statistical significance.

To further refine the feature set and reduce redundancy, Pearson's correlation coefficient was applied to assess collinearity among features. For pairs of features with high linear correlation (correlation coefficient > 0.9), only one was retained to minimize redundancy while preserving the integrity of the feature set. To further mitigate the risk of overfitting, we also employed the mRUS (minimum Redundancy Maximum Relevance) method, selecting the 32 most relevant features for the final model.

The remaining features were then input into a LASSO regression model, which penalizes the absolute values of the regression coefficients to facilitate feature selection (Supplementary Figure 4B). The optimal regularization parameter (λ) was determined through 10-fold cross-validation, selecting the value that minimized the cross-validation error (Supplementary Figure 4C). Only features with non-zero coefficients were retained, representing the most informative predictors (Supplementary Figure 4D). This rigorous feature selection and optimization process highlights the robustness of our approach and enhances the reliability of our findings.

**Construct radiomics models**

These features were used to construct predictive models using multiple machine learning algorithms (Supplementary Table 4). Similar to the MIL framework described above, the SMOTE method was applied during training to address class imbalance. Model robustness was evaluated using five-fold cross-validation on the training set, and hyperparameters were optimized through grid search.

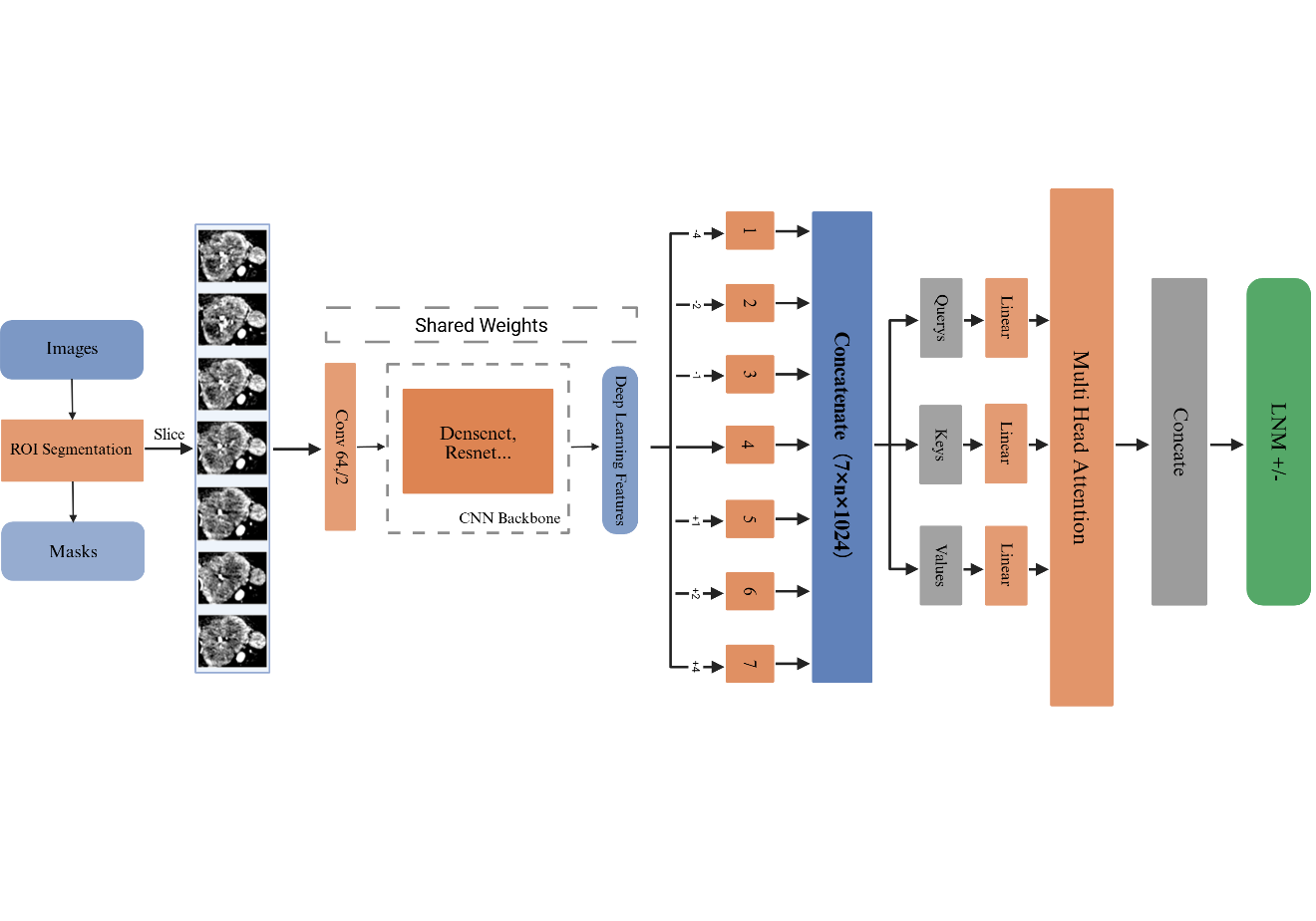
Supplementary Figure 1. The architecture of the Transformer model proposed by this study

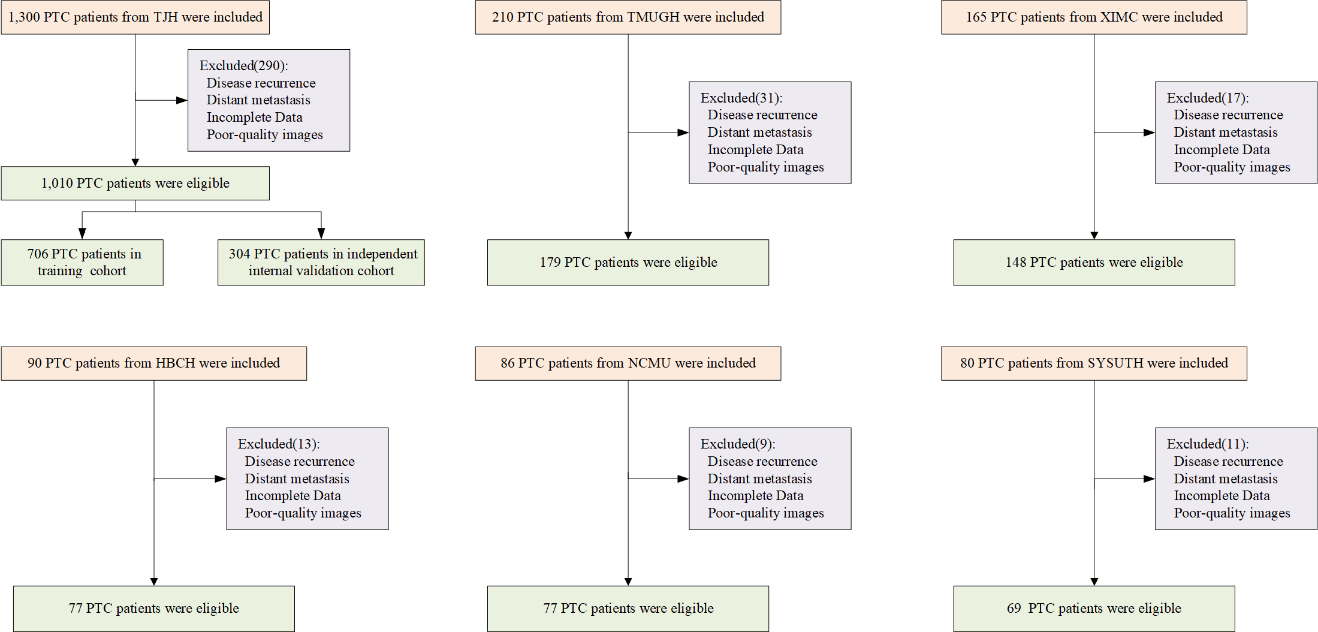

Supplementary Figure 2. Flowchart of patient selection for the training, validation, and external test cohorts.

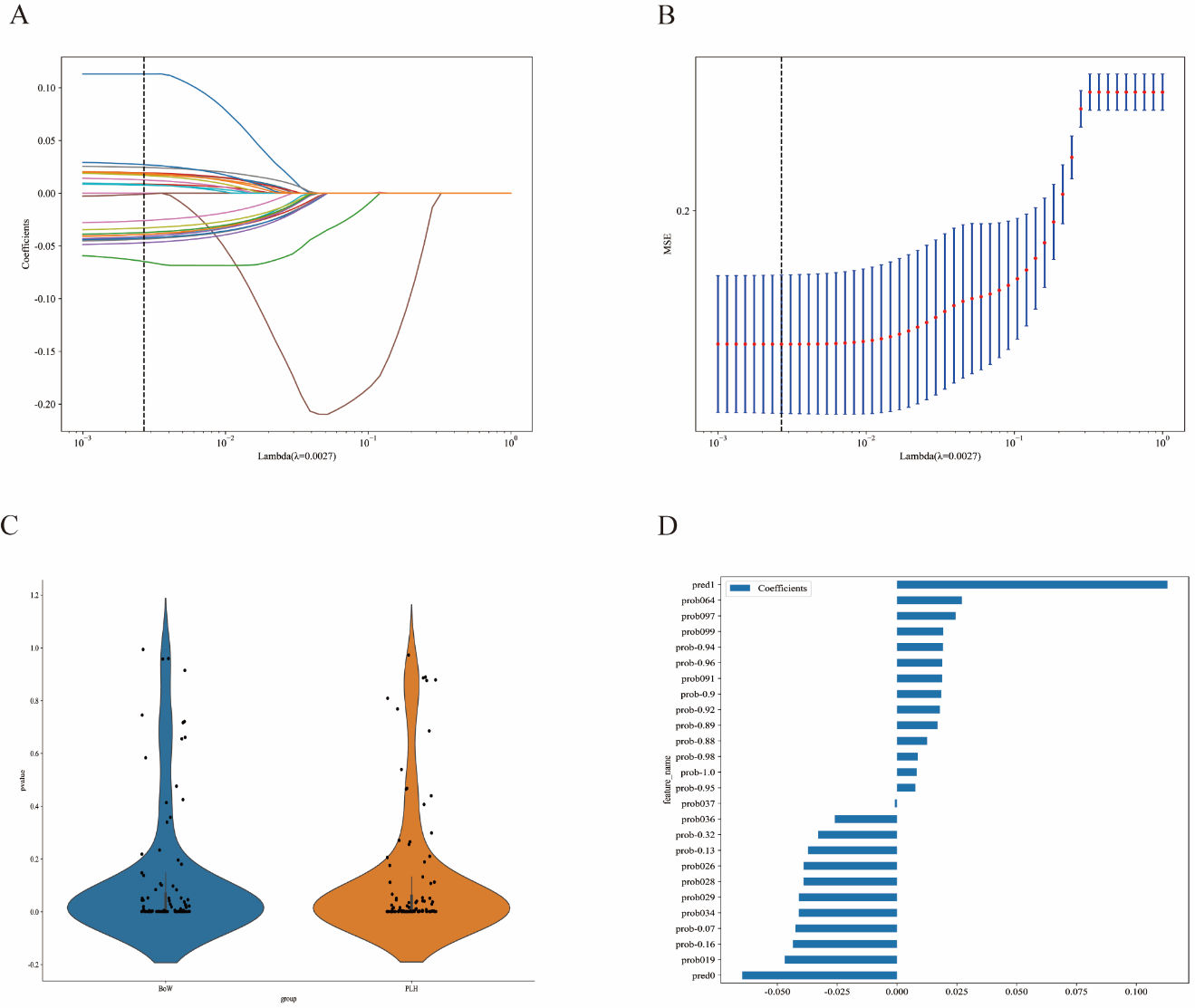
Supplementary Figure 3. (A) LASSO coefficient paths for MIL-derived fusion features (PLH and BoW) across different values of the regularization parameter (λ). (B) Five-fold cross-validation curve for selecting the optimal λ in LASSO. The dashed vertical line indicates the chosen λ (minimum cross-validated error). (C) Violin plots showing the distribution of MIL-derived feature representations summarized at the sample level using the BoW and PLH strategies. Each dot represents one sample. (D) Coefficients of the selected MIL fusion features retained in the final sparse model, indicating the direction and magnitude of each feature’s contribution.

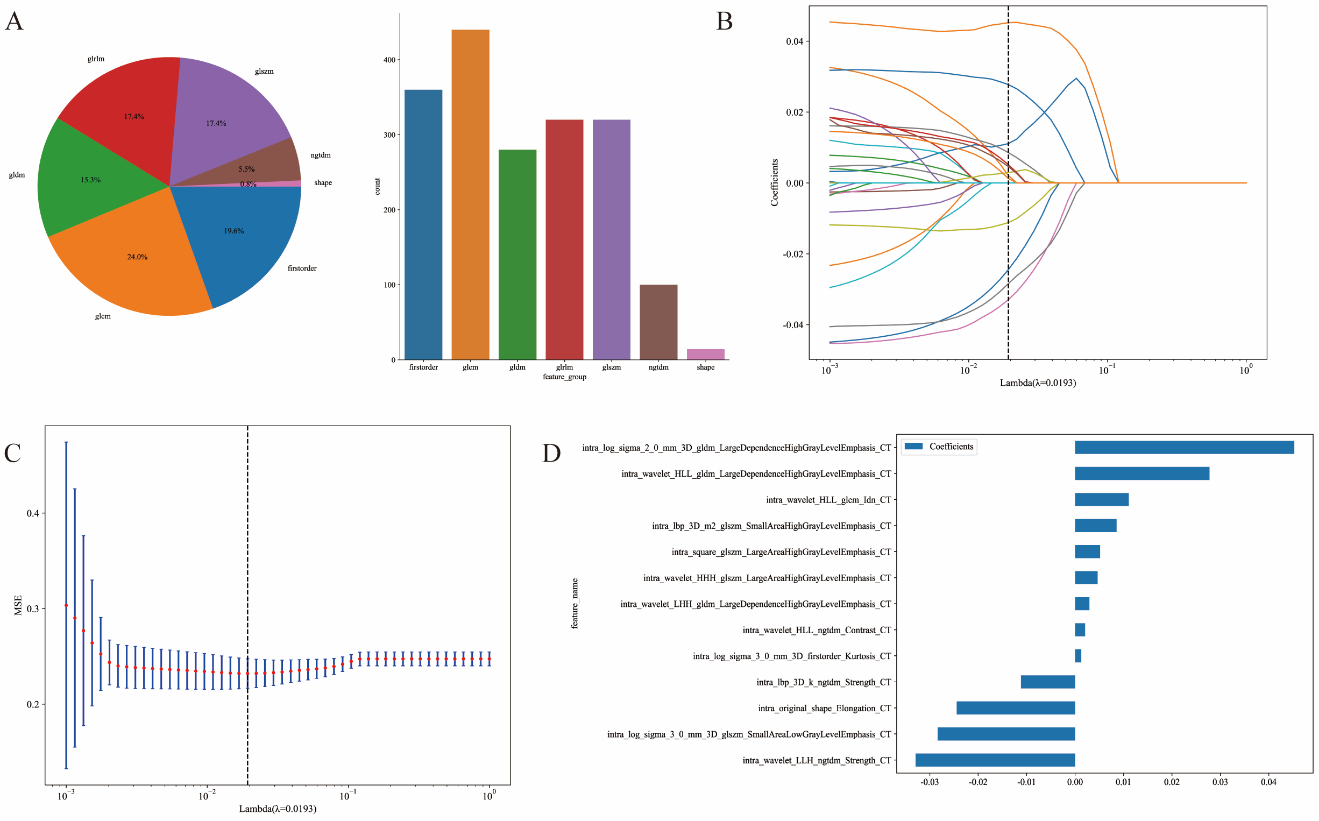

Supplementary Figure 4. (A) Distribution of extracted radiomic features by category and image transformation, including geometry, intensity, and texture features derived from original, LoG, and wavelet-transformed images. (B) LASSO coefficient profiles of radiomic features across different values of the regularization parameter (λ). (C) Ten-fold cross-validation curve for determining the optimal λ value in the LASSO model. The dashed vertical line indicates the selected λ corresponding to the minimum cross-validation error. (D) Non-zero coefficients of radiomic features retained in the final radiomics signature.

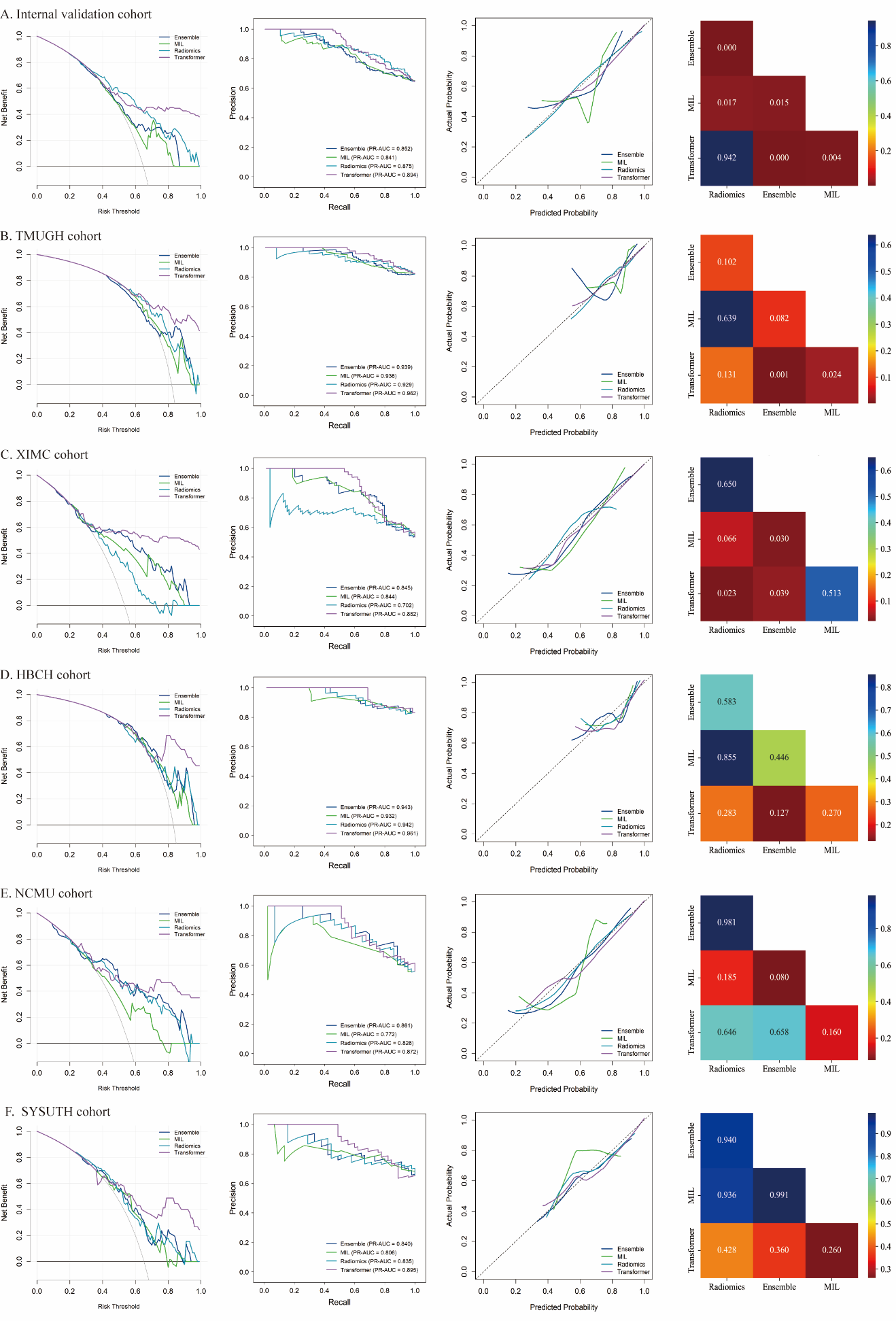

Supplementary Figure 5. Comparative performance of all models across the internal validation (A) and external cohorts (B–F). From left to right: decision curve analysis (DCA), precision–recall (PR) curves, calibration curves, and pairwise comparisons using the DeLong test.

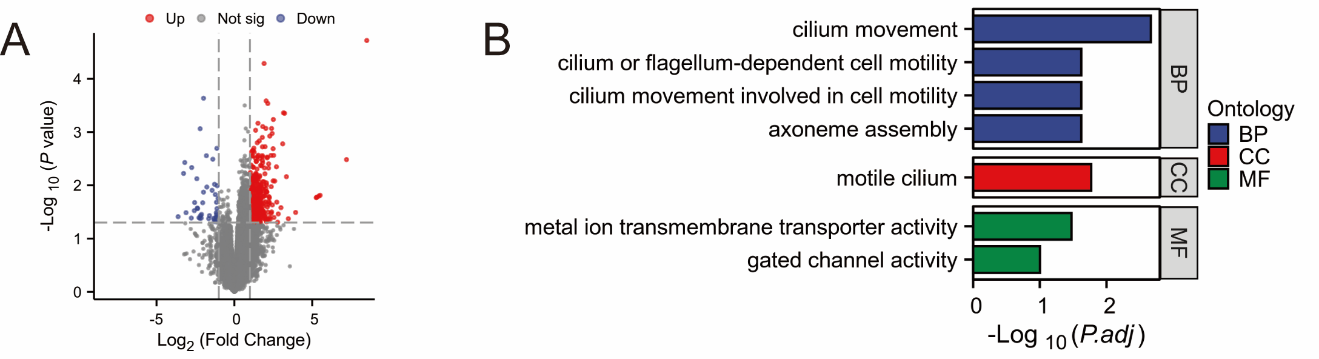

Supplementary Figure 6. (A) Volcano plot of DEGs between predicted groups. (B) GO and KEGG enrichment analysis of DEGs. Biological Process (BP), Cellular Component (CC), Molecular Function (MF), and KEGG pathway categories are shown.

Supplementary Table 1

| **Model** | **Accuracy** | **AUC (95% CI)** | **Sensitivity** | **Specificity** | **PPV** | **NPV** |
| --- | --- | --- | --- | --- | --- | --- |
| **Train Cohort** | | | | | | |
| resnet18 | 0.672 | 0.739 (0.725-0.753) | 0.662 | 0.685 | 0.727 | 0.616 |
| resnet50 | 0.650 | 0.708 (0.694-0.723) | 0.588 | 0.729 | 0.733 | 0.584 |
| resnet101 | 0.645 | 0.711 (0.697-0.725) | 0.602 | 0.699 | 0.716 | 0.582 |
| densenet121 | 0.662 | 0.732 (0.718-0.745) | 0.647 | 0.682 | 0.720 | 0.605 |
| densenet201 | 0.687 | 0.769 (0.756-0.782) | 0.647 | 0.737 | 0.756 | 0.623 |
| **Val Cohort** | | | | | | |
| resnet18 | 0.560 | 0.648 (0.625-0.671) | 0.420 | 0.818 | 0.810 | 0.434 |
| resnet50 | 0.569 | 0.642 (0.619-0.666) | 0.449 | 0.789 | 0.797 | 0.437 |
| resnet101 | 0.620 | 0.690 (0.667-0.713) | 0.518 | 0.808 | 0.832 | 0.476 |
| densenet121 | 0.611 | 0.695 (0.673-0.717) | 0.523 | 0.774 | 0.810 | 0.468 |
| densenet201 | 0.599 | 0.699 (0.677-0.721) | 0.450 | 0.873 | 0.867 | 0.463 |
| **TMUGH Cohort** | | | | | | |
| resnet18 | 0.600 | 0.702 (0.669-0.735) | 0.561 | 0.781 | 0.922 | 0.279 |
| resnet50 | 0.595 | 0.680 (0.644-0.716) | 0.562 | 0.750 | 0.912 | 0.271 |
| resnet101 | 0.638 | 0.719 (0.685-0.752) | 0.618 | 0.728 | 0.912 | 0.293 |
| densenet121 | 0.547 | 0.686 (0.652-0.720) | 0.491 | 0.808 | 0.922 | 0.257 |
| densenet201 | 0.595 | 0.702 (0.671-0.732) | 0.538 | 0.857 | 0.945 | 0.288 |
| **XIMC Cohort** | | | | | | |
| resnet18 | 0.643 | 0.682 (0.650-0.714) | 0.477 | 0.832 | 0.765 | 0.582 |
| resnet50 | 0.688 | 0.744 (0.715-0.774) | 0.606 | 0.783 | 0.761 | 0.634 |
| resnet101 | 0.712 | 0.764 (0.735-0.792) | 0.627 | 0.810 | 0.790 | 0.655 |
| densenet121 | 0.678 | 0.727 (0.696-0.757) | 0.613 | 0.752 | 0.739 | 0.629 |
| densenet201 | 0.702 | 0.759 (0.730-0.788) | 0.620 | 0.795 | 0.776 | 0.646 |
| **NCMU Cohort** | | | | | | |
| resnet18 | 0.601 | 0.609 (0.561-0.657) | 0.641 | 0.550 | 0.643 | 0.548 |
| resnet50 | 0.649 | 0.663 (0.617-0.709) | 0.757 | 0.513 | 0.663 | 0.626 |
| resnet101 | 0.653 | 0.714 (0.671-0.757) | 0.648 | 0.660 | 0.707 | 0.597 |
| densenet121 | 0.672 | 0.705 (0.661-0.749) | 0.621 | 0.735 | 0.748 | 0.606 |
| densenet201 | 0.690 | 0.756 (0.716-0.796) | 0.522 | 0.903 | 0.872 | 0.599 |
| **HBCH Cohort** | | | | | | |
| resnet18 | 0.636 | 0.640 (0.583-0.696) | 0.645 | 0.593 | 0.887 | 0.254 |
| resnet50 | 0.575 | 0.667 (0.608-0.725) | 0.538 | 0.758 | 0.916 | 0.250 |
| resnet101 | 0.622 | 0.698 (0.648-0.748) | 0.598 | 0.736 | 0.918 | 0.271 |
| densenet121 | 0.521 | 0.667 (0.616-0.718) | 0.458 | 0.835 | 0.932 | 0.238 |
| densenet201 | 0.662 | 0.739 (0.692-0.785) | 0.647 | 0.736 | 0.924 | 0.298 |
| **SYSUTH Cohort** | | | | | | |
| resnet18 | 0.627 | 0.681 (0.631-0.731) | 0.581 | 0.714 | 0.792 | 0.476 |
| resnet50 | 0.667 | 0.721 (0.674-0.767) | 0.648 | 0.702 | 0.803 | 0.515 |
| resnet101 | 0.673 | 0.716 (0.669-0.764) | 0.686 | 0.649 | 0.785 | 0.524 |
| densenet121 | 0.735 | 0.742 (0.696-0.788) | 0.838 | 0.542 | 0.774 | 0.641 |
| densenet201 | 0.588 | 0.694 (0.647-0.742) | 0.444 | 0.857 | 0.854 | 0.451 |

Supplementary Table 1. Performance metrics of different models for slice-level predictions

Supplementary Table 2

| Model | Accuracy | AUC (95% CI) | Sensitivity | Specificity | PPV | NPV |
| --- | --- | --- | --- | --- | --- | --- |
| **Train Cohort** | | | | | | |
| LR | 0.793 | 0.895 (0.873-0.917) | 0.784 | 0.804 | 0.835 | 0.747 |
| RF | 0.772 | 0.863 (0.838-0.889) | 0.708 | 0.853 | 0.858 | 0.698 |
| ET | 0.754 | 0.866 (0.840-0.891) | 0.673 | 0.856 | 0.855 | 0.674 |
| XGB | 0.775 | 0.862 (0.837-0.888) | 0.716 | 0.849 | 0.857 | 0.703 |
| LGBM | 0.779 | 0.874 (0.849-0.899) | 0.723 | 0.849 | 0.858 | 0.709 |
| **Val Cohort** | | | | | | |
| LR | 0.674 | 0.708 (0.650-0.767) | 0.838 | 0.374 | 0.711 | 0.556 |
| RF | 0.645 | 0.715 (0.658-0.772) | 0.792 | 0.374 | 0.700 | 0.494 |
| ET | 0.651 | 0.706 (0.647-0.766) | 0.792 | 0.393 | 0.706 | 0.506 |
| XGB | 0.645 | 0.702 (0.644-0.760) | 0.792 | 0.374 | 0.700 | 0.494 |
| LGBM | 0.651 | 0.700 (0.641-0.759) | 0.797 | 0.383 | 0.704 | 0.506 |
| **TMUGH Cohort** | | | | | | |
| LR | 0.709 | 0.723 (0.644-0.803) | 0.762 | 0.469 | 0.868 | 0.300 |
| RF | 0.693 | 0.730 (0.652-0.808) | 0.735 | 0.500 | 0.871 | 0.291 |
| ET | 0.687 | 0.725 (0.642-0.808) | 0.721 | 0.531 | 0.876 | 0.293 |
| XGB | 0.698 | 0.709 (0.625-0.794) | 0.741 | 0.500 | 0.872 | 0.296 |
| LGBM | 0.693 | 0.716 (0.632-0.799) | 0.735 | 0.500 | 0.871 | 0.291 |
| **XIMC Cohort** | | | | | | |
| LR | 0.736 | 0.797 (0.723-0.870) | 0.785 | 0.681 | 0.738 | 0.734 |
| RF | 0.709 | 0.793 (0.722-0.864) | 0.734 | 0.681 | 0.725 | 0.691 |
| ET | 0.723 | 0.794 (0.720-0.867) | 0.734 | 0.710 | 0.744 | 0.700 |
| XGB | 0.696 | 0.780 (0.706-0.854) | 0.734 | 0.652 | 0.707 | 0.682 |
| LGBM | 0.703 | 0.782 (0.707-0.856) | 0.734 | 0.667 | 0.716 | 0.687 |
| **NCMU Cohort** | | | | | | |
| LR | 0.636 | 0.729 (0.617-0.842) | 0.884 | 0.324 | 0.623 | 0.687 |
| RF | 0.623 | 0.722 (0.610-0.834) | 0.884 | 0.294 | 0.613 | 0.667 |
| ET | 0.636 | 0.727 (0.616-0.839) | 0.884 | 0.324 | 0.623 | 0.687 |
| XGB | 0.623 | 0.710 (0.596-0.824) | 0.884 | 0.294 | 0.613 | 0.667 |
| LGBM | 0.623 | 0.706 (0.590-0.822) | 0.884 | 0.294 | 0.613 | 0.667 |
| **HBCH Cohort** | | | | | | |
| LR | 0.740 | 0.775 (0.661-0.890) | 0.781 | 0.538 | 0.893 | 0.333 |
| RF | 0.740 | 0.721 (0.578-0.863) | 0.797 | 0.462 | 0.879 | 0.316 |
| ET | 0.740 | 0.743 (0.618-0.868) | 0.797 | 0.462 | 0.879 | 0.316 |
| XGB | 0.740 | 0.704 (0.554-0.854) | 0.797 | 0.462 | 0.879 | 0.316 |
| LGBM | 0.740 | 0.713 (0.561-0.864) | 0.797 | 0.462 | 0.879 | 0.316 |
| **SYSUTH Cohort** | | | | | | |
| LR | 0.681 | 0.685 (0.551-0.818) | 0.711 | 0.625 | 0.780 | 0.536 |
| RF | 0.652 | 0.712 (0.582-0.842) | 0.667 | 0.625 | 0.769 | 0.500 |
| ET | 0.652 | 0.694 (0.567-0.821) | 0.667 | 0.625 | 0.769 | 0.500 |
| XGB | 0.652 | 0.694 (0.558-0.831) | 0.667 | 0.625 | 0.769 | 0.500 |
| LGBM | 0.652 | 0.695 (0.560-0.830) | 0.667 | 0.625 | 0.769 | 0.500 |

Supplementary Table 2. Performance metrics of different models for MIL fusion

Supplementary Table 3

| **Method** | **Accuracy** | **AUC (95% CI)** | **Sensitivity** | **Specificity** | **PPV** | **NPV** |
| --- | --- | --- | --- | --- | --- | --- |
| **Train Cohort** | | | | | | |
| mean | 0.800 | 0.895 (0.873-0.917) | 0.845 | 0.744 | 0.806 | 0.792 |
| max | 0.599 | 0.835 (0.806-0.864) | 0.992 | 0.103 | 0.583 | 0.914 |
| **Val Cohort** | | | | | | |
| mean | 0.661 | 0.718 (0.662-0.774) | 0.868 | 0.280 | 0.690 | 0.536 |
| max | 0.658 | 0.726 (0.670-0.781) | 0.970 | 0.084 | 0.661 | 0.600 |
| **TMUGH Cohort** | | | | | | |
| mean | 0.732 | 0.738 (0.662-0.814) | 0.789 | 0.469 | 0.872 | 0.326 |
| max | 0.793 | 0.776 (0.698-0.853) | 0.932 | 0.156 | 0.835 | 0.333 |
| **XIMC Cohort** | | | | | | |
| mean | 0.723 | 0.794 (0.721-0.868) | 0.797 | 0.638 | 0.716 | 0.733 |
| max | 0.628 | 0.810 (0.742-0.879) | 0.962 | 0.246 | 0.594 | 0.850 |
| **NCMU Cohort** | | | | | | |
| mean | 0.662 | 0.799 (0.698-0.899) | 0.884 | 0.382 | 0.644 | 0.722 |
| max | 0.558 | 0.766 (0.659-0.873) | 0.977 | 0.029 | 0.560 | 0.500 |
| **HBCH Cohort** | | | | | | |
| mean | 0.740 | 0.744 (0.618-0.870) | 0.812 | 0.385 | 0.867 | 0.294 |
| max | 0.844 | 0.780 (0.663-0.897) | 0.953 | 0.308 | 0.871 | 0.571 |
| **SYSUTH Cohort** | | | | | | |
| mean | 0.696 | 0.726 (0.600-0.851) | 0.733 | 0.625 | 0.786 | 0.556 |
| max | 0.696 | 0.747 (0.624-0.869) | 0.978 | 0.167 | 0.687 | 0.800 |

Supplementary Table 3. Performance metrics of different methods for ensemble fusion

Supplementary Table 4

| **Model** | **Accuracy** | **AUC (95% CI)** | **Sensitivity** | **Specificity** | **PPV** | **NPV** |
| --- | --- | --- | --- | --- | --- | --- |
| **Train Cohort** | | | | | | |
| LR | 0.618 | 0.677 (0.637-0.716) | 0.581 | 0.663 | 0.686 | 0.556 |
| SVM | 0.636 | 0.738 (0.701-0.774) | 0.551 | 0.744 | 0.731 | 0.567 |
| RF | 0.708 | 0.802 (0.770-0.834) | 0.629 | 0.808 | 0.805 | 0.633 |
| ET | 0.629 | 0.720 (0.682-0.757) | 0.566 | 0.708 | 0.710 | 0.564 |
| XGB | 0.661 | 0.733 (0.698-0.768) | 0.528 | 0.830 | 0.797 | 0.582 |
| LGBM | 0.653 | 0.726 (0.689-0.762) | 0.558 | 0.772 | 0.756 | 0.581 |
| **Val Cohort** | | | | | | |
| LR | 0.674 | 0.776 (0.724-0.829) | 0.589 | 0.832 | 0.866 | 0.524 |
| SVM | 0.612 | 0.717 (0.659-0.775) | 0.497 | 0.822 | 0.838 | 0.471 |
| RF | 0.664 | 0.785 (0.733-0.837) | 0.563 | 0.850 | 0.874 | 0.514 |
| ET | 0.674 | 0.779 (0.728-0.831) | 0.579 | 0.850 | 0.877 | 0.523 |
| XGB | 0.599 | 0.725 (0.671-0.779) | 0.457 | 0.860 | 0.857 | 0.462 |
| LGBM | 0.628 | 0.756 (0.702-0.810) | 0.497 | 0.869 | 0.875 | 0.484 |
| **TMUGH Cohort** | | | | | | |
| LR | 0.609 | 0.751 (0.660-0.843) | 0.571 | 0.781 | 0.923 | 0.284 |
| SVM | 0.497 | 0.670 (0.568-0.773) | 0.435 | 0.781 | 0.901 | 0.231 |
| RF | 0.570 | 0.752 (0.661-0.842) | 0.510 | 0.844 | 0.937 | 0.273 |
| ET | 0.564 | 0.748 (0.654-0.842) | 0.510 | 0.812 | 0.926 | 0.265 |
| XGB | 0.503 | 0.672 (0.574-0.770) | 0.442 | 0.781 | 0.903 | 0.234 |
| LGBM | 0.559 | 0.735 (0.647-0.824) | 0.503 | 0.812 | 0.925 | 0.263 |
| **XIMC Cohort** | | | | | | |
| LR | 0.649 | 0.694 (0.608-0.781) | 0.671 | 0.623 | 0.671 | 0.623 |
| SVM | 0.588 | 0.682 (0.597-0.768) | 0.557 | 0.623 | 0.629 | 0.551 |
| RF | 0.662 | 0.697 (0.612-0.783) | 0.684 | 0.638 | 0.684 | 0.638 |
| ET | 0.669 | 0.692 (0.605-0.778) | 0.696 | 0.638 | 0.687 | 0.647 |
| XGB | 0.601 | 0.685 (0.599-0.770) | 0.456 | 0.768 | 0.692 | 0.552 |
| LGBM | 0.615 | 0.677 (0.590-0.764) | 0.570 | 0.667 | 0.662 | 0.575 |
| **NCMU Cohort** | | | | | | |
| LR | 0.753 | 0.795 (0.694-0.897) | 0.674 | 0.853 | 0.853 | 0.674 |
| SVM | 0.727 | 0.789 (0.685-0.892) | 0.581 | 0.912 | 0.893 | 0.633 |
| RF | 0.701 | 0.778 (0.673-0.884) | 0.651 | 0.765 | 0.778 | 0.634 |
| ET | 0.727 | 0.793 (0.691-0.895) | 0.628 | 0.853 | 0.844 | 0.644 |
| XGB | 0.714 | 0.776 (0.671-0.882) | 0.674 | 0.765 | 0.784 | 0.650 |
| LGBM | 0.688 | 0.779 (0.677-0.882) | 0.628 | 0.765 | 0.771 | 0.619 |
| **HBCH Cohort** | | | | | | |
| LR | 0.688 | 0.756 (0.628-0.884) | 0.703 | 0.615 | 0.900 | 0.296 |
| SVM | 0.675 | 0.772 (0.642-0.903) | 0.656 | 0.769 | 0.933 | 0.312 |
| RF | 0.649 | 0.736 (0.614-0.857) | 0.672 | 0.538 | 0.878 | 0.250 |
| ET | 0.649 | 0.714 (0.576-0.852) | 0.672 | 0.538 | 0.878 | 0.250 |
| XGB | 0.584 | 0.581 (0.400-0.761) | 0.594 | 0.538 | 0.864 | 0.212 |
| LGBM | 0.597 | 0.696 (0.560-0.832) | 0.594 | 0.615 | 0.884 | 0.235 |
| **SYSUTH Cohort** | | | | | | |
| LR | 0.536 | 0.707 (0.575-0.840) | 0.378 | 0.833 | 0.810 | 0.417 |
| SVM | 0.507 | 0.619 (0.481-0.757) | 0.333 | 0.833 | 0.789 | 0.400 |
| RF | 0.594 | 0.718 (0.591-0.844) | 0.422 | 0.917 | 0.905 | 0.458 |
| ET | 0.522 | 0.700 (0.572-0.828) | 0.356 | 0.833 | 0.800 | 0.408 |
| XGB | 0.594 | 0.678 (0.555-0.801) | 0.444 | 0.875 | 0.870 | 0.457 |
| LGBM | 0.594 | 0.671 (0.537-0.805) | 0.444 | 0.875 | 0.870 | 0.457 |

Supplementary Table 4. Performance metrics of traditional radiomics models

Supplementary Table 5

|  | **AUC (95% CI)** | ***P* value** | **Sensitivity** | **Specificity** | **PPV** | **NPV** |
| --- | --- | --- | --- | --- | --- | --- |
| **Train cohort** |  |  |  |  |  |  |
| ThyLNT | 0.692 (0.660-0.723) | -- | 0.896 | 0.487 | 0.688 | 0.788 |
| US | 0.656 (0.621-0.690) | <0.001 | 0.571 | 0.740 | 0.735 | 0.578 |
| CT | 0.624 (0.588-0.659) | <0.001 | 0.584 | 0.664 | 0.687 | 0.558 |
| US+CT | 0.638 (0.602-0.673) | <0.001 | 0.721 | 0.555 | 0.671 | 0.611 |
| ThyLNT+US | 0.888 (0.864-0.911) | 0.237 | 0.782 | 0.859 | 0.875 | 0.757 |
| ThyLNT+CT | 0.883 (0.858-0.907) | 0.841 | 0.774 | 0.875 | 0.887 | 0.754 |
| ThyLNT+US+CT | 0.888 (0.864-0.911) | 0.254 | 0.797 | 0.849 | 0.870 | 0.768 |
| **Internal validation cohort** | |  |  |  |  |  |
| ThyLNT | 0.659 (0.605-0.714) | -- | 0.833 | 0.486 | 0.749 | 0.612 |
| US | 0.655 (0.600-0.710) | <0.001 | 0.599 | 0.710 | 0.792 | 0.490 |
| CT | 0.621 (0.564-0.678) | <0.001 | 0.579 | 0.664 | 0.760 | 0.461 |
| US+CT | 0.617 (0.560-0.674) | <0.001 | 0.721 | 0.514 | 0.732 | 0.500 |
| ThyLNT+US | 0.803 (0.745-0.851) | 0.459 | 0.173 | 1.000 | 1.000 | 0.396 |
| ThyLNT+CT | 0.791 (0.741-0.840) | 0.855 | 0.168 | 1.000 | 1.000 | 0.395 |
| ThyLNT+US+CT | 0.794 (0.745-0.843) | 0.779 | 0.188 | 1.000 | 1.000 | 0.401 |
| **TMUGH cohort** |  |  |  |  |  |  |
| ThyLNT | 0.758 (0.669-0.847) | -- | 0.891 | 0.625 | 0.916 | 0.556 |
| US | 0.768 (0.710-0.826) | 0.186 | 0.599 | 0.938 | 0.978 | 0.337 |
| CT | 0.617 (0.538-0.697) | <0.001 | 0.422 | 0.813 | 0.912 | 0.234 |
| US+CT | 0.729 (0.644-0.813) | 0.088 | 0.708 | 0.750 | 0.929 | 0.358 |
| ThyLNT+US | 0.891 (0.842-0.940) | 0.018 | 0.116 | 1.000 | 1.000 | 0.198 |
| ThyLNT+CT | 0.809 (0.734-0.884) | 0.652 | 0.109 | 1.000 | 1.000 | 0.196 |
| ThyLNT+US+CT | 0.876 (0.823-0.929) | 0.177 | 0.116 | 1.000 | 1.000 | 0.198 |
| **XIMC cohort** |  |  |  |  |  |  |
| ThyLNT | 0.671 (0.596-0.747) | -- | 0.734 | 0.609 | 0.682 | 0.667 |
| US | 0.546 (0.479-0.613) | <0.001 | 0.266 | 0.826 | 0.636 | 0.496 |
| CT | 0.497 (0.427-0.567) | <0.001 | 0.241 | 0.754 | 0.528 | 0.464 |
| US+CT | 0.516 (0.438-0.594) | <0.001 | 0.380 | 0.652 | 0.556 | 0.479 |
| ThyLNT+US | 0.759 (0.682-0.836) | 0.054 | 0.127 | 1.000 | 1.000 | 0.500 |
| ThyLNT+CT | 0.751 (0.673-0.828) | 0.036 | 0.114 | 1.000 | 1.000 | 0.496 |
| ThyLNT+US+CT | 0.725 (0.645-0.806) | 0.017 | 0.114 | 1.000 | 1.000 | 0.496 |
| **HBCH cohort** |  |  |  |  |  |  |
| ThyLNT | 0.569 (0.466-0.672) | -- | 0.984 | 0.154 | 0.851 | 0.667 |
| US | 0.591 (0.455-0.728) | 0.032 | 0.875 | 0.308 | 0.862 | 0.333 |
| CT | 0.623 (0.489-0.757) | 0.073 | 0.938 | 0.308 | 0.870 | 0.500 |
| US+CT | 0.576 (0.453-0.700) | 0.015 | 0.922 | 0.231 | 0.855 | 0.375 |
| ThyLNT+US | 0.823 (0.724-0.922) | 0.711 | 0.109 | 1.000 | 1.000 | 0.186 |
| ThyLNT+CT | 0.864 (0.772-0.956) | 0.264 | 0.078 | 1.000 | 1.000 | 0.181 |
| ThyLNT+US+CT | 0.839 (0.743-0.935) | 0.577 | 0.109 | 1.000 | 1.000 | 0.186 |
| **NCMU cohort** |  |  |  |  |  |  |
| ThyLNT | 0.816 (0.733-0.900) | -- | 0.721 | 0.912 | 0.912 | 0.721 |
| US | 0.680 (0.595-0.765) | 0.039 | 0.419 | 0.941 | 0.900 | 0.561 |
| CT | 0.660 (0.582-0.737) | 0.014 | 0.349 | 0.971 | 0.938 | 0.541 |
| US+CT | 0.723 (0.634-0.813) | 0.179 | 0.535 | 0.912 | 0.885 | 0.608 |
| ThyLNT+US | 0.836 (0.747-0.925) | 0.186 | 0.047 | 1.000 | 1.000 | 0.453 |
| ThyLNT+CT | 0.831 (0.741-0.921) | 0.180 | 0.047 | 1.000 | 1.000 | 0.453 |
| ThyLNT+US+CT | 0.842 (0.755-0.929) | 0.160 | 0.047 | 1.000 | 1.000 | 0.453 |
| **SYSUTH cohort** |  |  |  |  |  |  |
| ThyLNT | 0.782 (0.686-0.878) | -- | 0.689 | 0.875 | 0.912 | 0.600 |
| US | 0.622 (0.559-0.686) | 0.017 | 0.244 | 1.000 | 1.000 | 0.414 |
| CT | 0.649 (0.549-0.748) | 0.075 | 0.422 | 0.875 | 0.864 | 0.447 |
| US+CT | 0.660 (0.560-0.760) | 0.112 | 0.444 | 0.875 | 0.870 | 0.457 |
| ThyLNT+US | 0.816 (0.716-0.916) | 0.097 | 0.111 | 1.000 | 1.000 | 0.375 |
| ThyLNT+CT | 0.819 (0.716-0.921) | 0.269 | 0.089 | 1.000 | 1.000 | 0.369 |
| ThyLNT+US+CT | 0.838 (0.742-0.933) | 0.132 | 0.111 | 1.000 | 1.000 | 0.375 |

Supplementary Table 5. Performance metrics of ThyLNT and thyroid imaging across all cohorts
